# Identification of shared and differentiating genetic risk for autism spectrum disorder, attention deficit hyperactivity disorder and case subgroups

**DOI:** 10.1101/2021.05.20.21257484

**Authors:** Manuel Mattheisen, Jakob Grove, Thomas D Als, Joanna Martin, Georgios Voloudakis, Sandra Meier, Ditte Demontis, Jaroslav Bendl, Raymond Walters, Caitlin E Carey, Anders Rosengren, Nora Strom, Mads Engel Hauberg, Biao Zeng, Gabriel Hoffman, Jonas Bybjerg-Grauholm, Marie Bækvad-Hansen, Esben Agerbo, Bru Cormand, Merete Nordentoft, Thomas Werge, Ole Mors, David M Hougaard, Joseph D Buxbaum, Stephen V Faraone, Barbara Franke, Søren Dalsgaard, Preben B Mortensen, Elise B Robinson, Panos Roussos, Benjamin M Neale, Mark J Daly, Anders D Børglum

## Abstract

Attention deficit hyperactivity disorder (ADHD) and autism spectrum disorder (ASD) are highly heritable neurodevelopmental disorders with a considerable overlap in their genetic etiology. We dissected their shared and distinct genetic architecture by cross-disorder analyses of large data sets, including samples with information on comorbid diagnoses. We identified seven loci shared by the disorders and the first five genome-wide significant loci differentiating the disorders. All five differentiating loci showed opposite allelic directions in the two disorders separately as well as significant associations with variation in other traits e.g. educational attainment, items of neuroticism and regional brain volume. Integration with brain transcriptome data identified and prioritized several significantly associated genes. Genetic correlation of the shared liability across ASD-ADHD was strong for other psychiatric phenotypes while the ASD-ADHD differentiating liability correlated most strongly with cognitive traits. Polygenic score analyses revealed that individuals diagnosed with both ASD and ADHD are double-burdened with genetic risk for both disorders and show distinctive patterns of genetic association with other traits when compared to the ASD-only and ADHD-only subgroups. The results provide novel insights into the biological foundation for developing just one or both of the disorders and for driving the psychopathology discriminatively towards either ADHD or ASD.

## Introduction

Attention deficit hyperactivity disorder (ADHD) and autism spectrum disorder (ASD) are among the most common neurodevelopmental disorders in children and often persist throughout adulthood ^1^. ADHD and ASD are both highly heritable (60-93%) ^2-4^ and the mode of their inheritance is complex and polygenic. Despite the high family-based heritability estimates, genome-wide association studies (GWAS) have only recently identified common variants robustly associated with each disorder ^5-7^. Although differing from one another with regard to core clinical symptoms, genetic studies have demonstrated significant overlap between the two disorders, with a genetic correlation (*r*_*G*_) from common variation of 0.36 ^5,8^ and substantial sharing of rare genetic risk variants such as large copy number variants ^9^ and protein-truncating variants ^10^. These findings are consistent with clinical and epidemiological evidence showing overlap in phenotypic features ^11^, high comorbidity rates between ASD and ADHD ^12,13^ in both females and males ^14^, and familial co-aggregation of the disorders with increased risk of ADHD among relatives of ASD probands (odds ratios monozygotic twins: 17.8, dizygotic twins: 4.3; full-siblings: 4.6, full cousins: 1.6) ^15^. Identification of the genetic components that are shared or distinct for the disorders may provide insights into the underlying biology and potentially inform on sub-classification, course and treatment.

Here, we utilize large collections of genotyped samples of ADHD and ASD from the Psychiatric Genomics Consortium (PGC) and the Lundbeck Foundation Initiative for Integrative Psychiatric Research (iPSYCH) to address two questions: 1) What specific variants and genes are shared by, or differentiate, ASD and ADHD? 2) Are there distinct genetic signatures in terms of polygenic burden for subgroups within these disorders such as cases diagnosed with both disorders (comorbid cases) or with just one of them (ASD-only, ADHD-only cases)?

## Results

### Shared genetic liability to ADHD and ASD

We performed a GWAS of diagnosed ADHD and/or ASD combined into a single phenotype (“combined GWAS”), totaling 34,462 cases and 41,201 controls on 8.9 million SNP allele dosages imputed from 1000 genomes phase 3 ^16^. Using LD score regression (LDSC) ^17^ we found evidence for a strong polygenic signal for this GWAS with an intercept of 1.0134 (ratio = 0.0558) and calculated the liability scale SNP-heritability to be 0.128 (for an assumed population prevalence of 0.055). We identified 263 genome-wide significant SNPs in seven distinct loci (**Table 1, Figure 1, Supplemental Figure S1**). All these loci showed associations with both of the disorders separately at p-values below 1×10^−4^ except one, which is genome-wide significant in ADHD and has a p-value of 0.009 in ASD. Overall, the findings corroborate previous results ^8,18^, but two loci have not been identified before as shared between ADHD and ASD. The novel shared associations are located in a highly pleiotropic multigene locus on chromosome 1 (rs7538463) and on chromosome 4 (rs227293) in *MANBA* (which encodes beta-D-mannoside mannohydrolase). Mutations in *MANBA* are associated with beta-mannosidosis, a lysosomal storage disease that has a wide spectrum of neurological phenotypes, including intellectual disability, hearing loss and speech impairment ^19^. More details on the seven loci can be found in **Table 1** and results from lookups in the open GWAS project database (https://gwas.mrcieu.ac.uk/about/, accessed Oct 14^th^ 2020) are available in **Supplemental Table S1** and as PheWAS plots in **Supplemental Figure S2**.

**Table 1:**
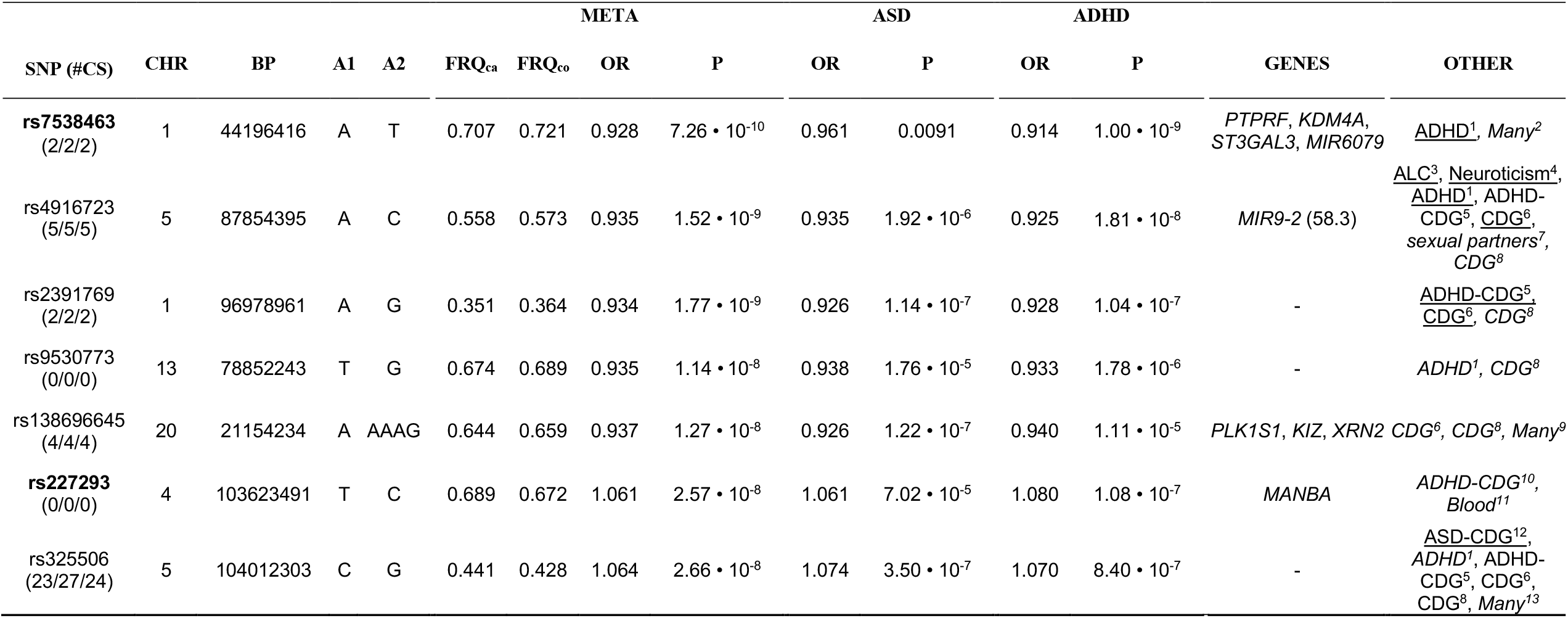
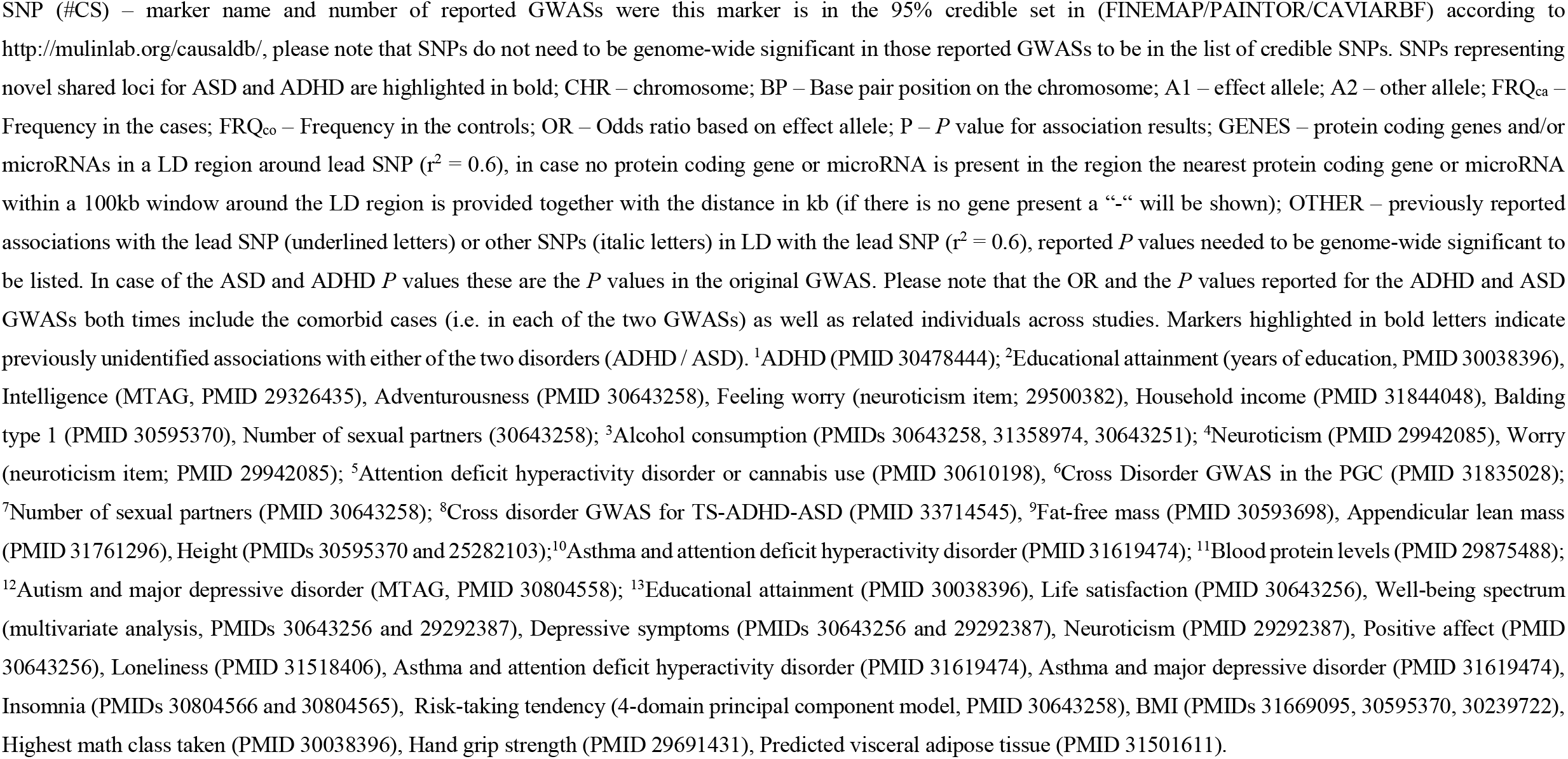
Results of combined (*ADHD or ASD*) GWAS.

**Figure 1:**
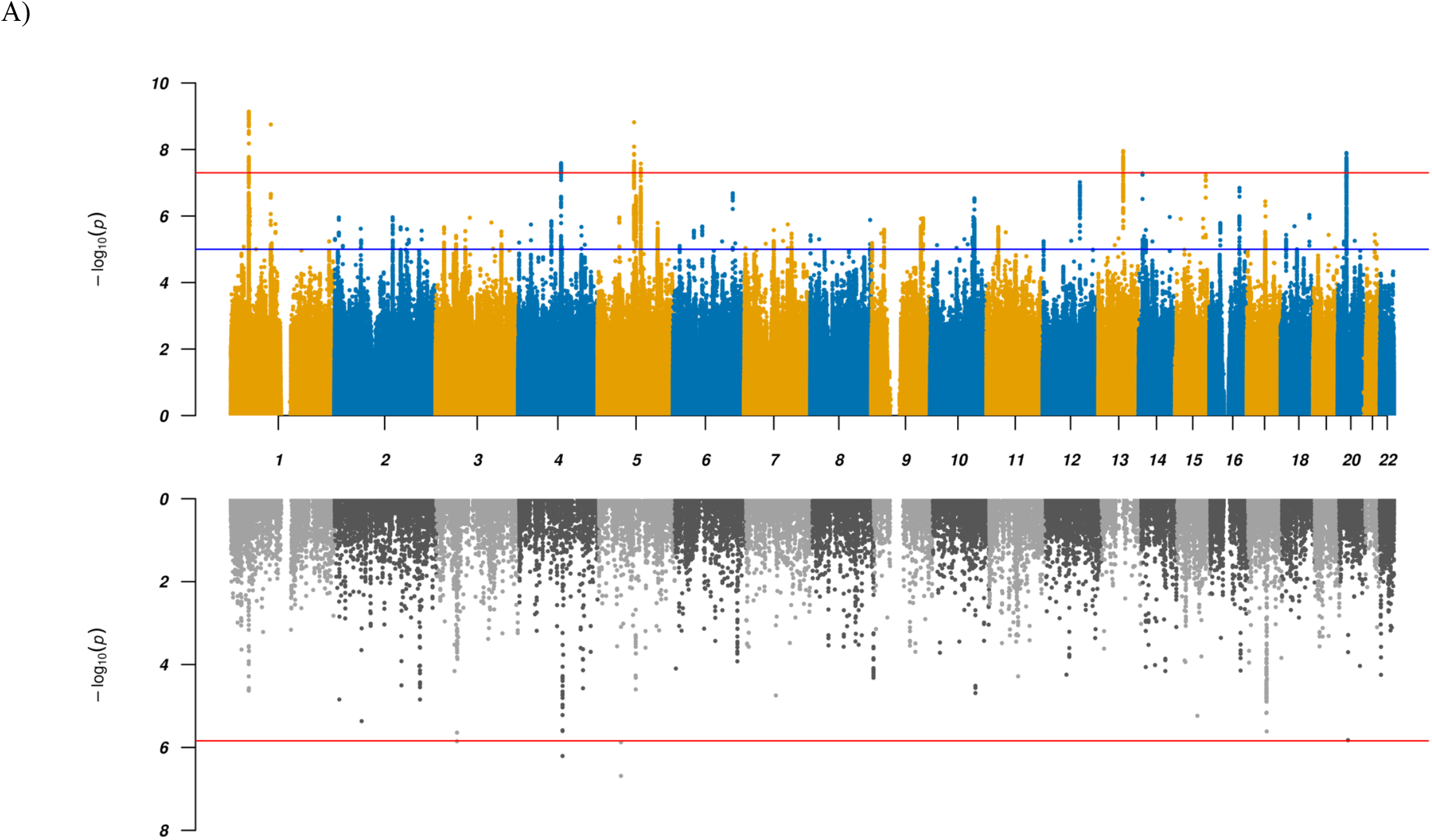
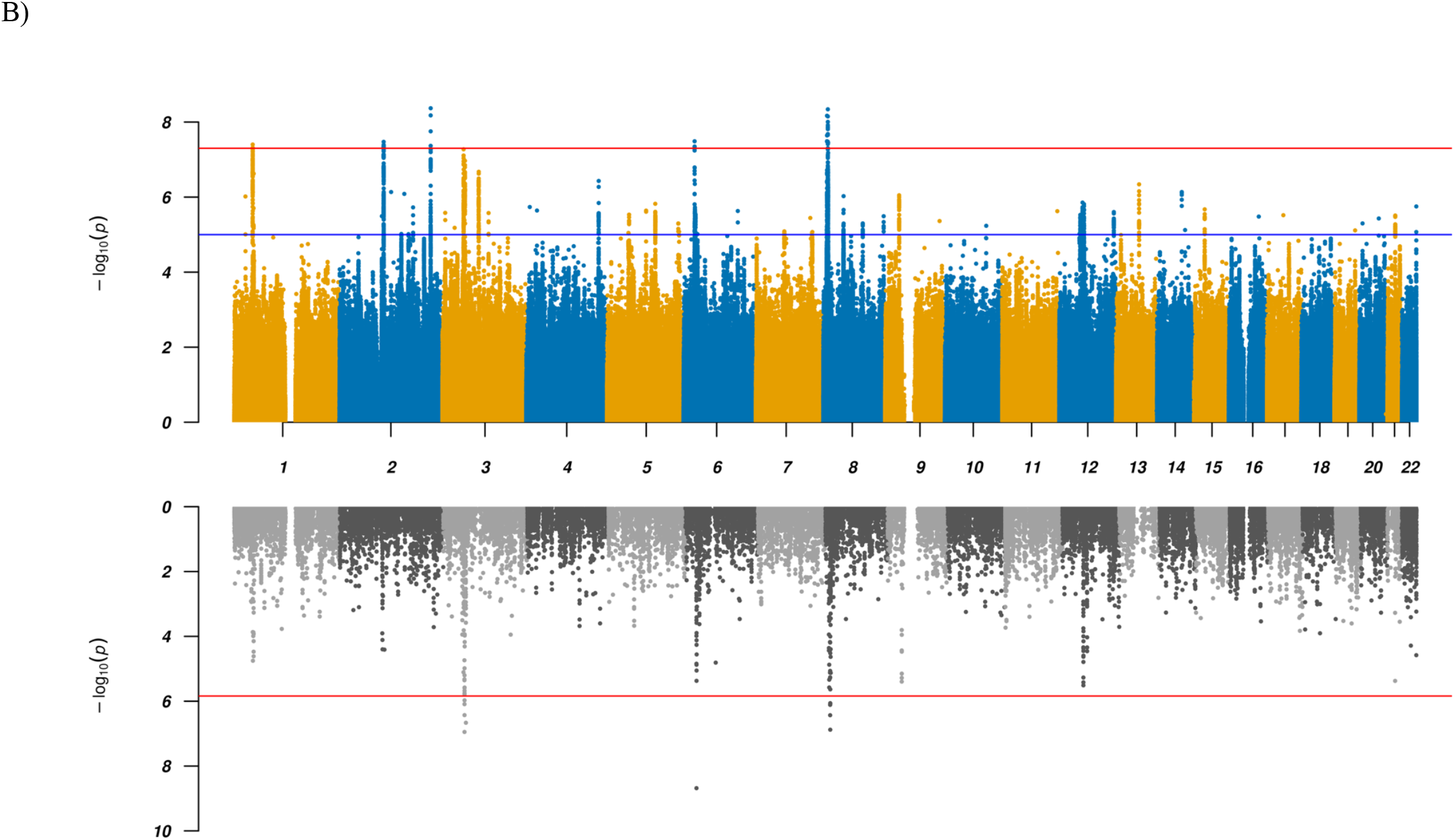
Miami plots of GWAS and TWAS results. Results for GWAS (top panels) and TWAS results for DLPFC transcripts (bottom panels) for (A) combined and (B) ADHD vs ASD GWAS. In the top panel a blue line in the Manhattan plot indicates a p-value of 1×10^−5^, a red line a p-value of 5×10^−8^ (genome-wide significance). Each dot represents a tested SNP. In the bottom panel genes are represented by both gene expression and isoform expression (= features, represented by the dots). A red line indicates Bonferroni corrected genome-wide significance within analyses (combined or ADHD vs ASD; p < 1.44×10^−6^; corresponding to Bonferroni correction of all the 34,646 features).

To identify and prioritize putative causal shared genes we performed a transcriptome-wide association study (TWAS), imputing the genetically regulated gene expression using EpiXcan ^20^ and expression data from the PsychENCODE Consortium ^21^ for genes as well as isoforms detected in 924 samples from the dorsolateral prefrontal cortex (DLPFC). Applying a conservative significance threshold (*p* < 1.44×10^−6^; corresponding to Bonferroni correction of all the 34,646 genes and isoforms tested), we identified five genes/isoforms showing significant differential expression between the combined case group and controls, and 177 genes/isoforms significant at a false discovery rate (FDR) < 0.05 (**Supplemental Table S2** and **Figure 1**). One of the five Bonferroni significant transcripts, the *KRT8P46-201* isoform, is located in the identified chromosome 4 GWAS locus in an intron of *MANBA* (which itself is among the genes with an FDR < 0.05) as illustrated in **Supplemental Figure S3a**. The four other top findings are the two genes *MOCS2* and *CCDC71* or their isoforms, which are not located in any of the identified GWAS loci and thus represent additional novel candidate genes for shared ADHD and ASD risk.

Gene-based analysis using MAGMA v 1.08 ^22^ with default settings as implemented in FUMA ^23^ largely corroborated the results from the GWAS and TWAS, highlighting e.g. *MANBA* (**Supplemental Figure S4a** and **Supplemental Table S3**). Furthermore, two of the significant genes (*SORCS3* and *DUSP6*) are located in regions that were not identified in the GWAS, suggesting these as additional shared loci.

### Differentiating genetic liability to ADHD and ASD

To identify loci with divergent effects on ADHD and ASD, we performed an association analysis comparing 11,964 ADHD-only cases with 9,315 ASD-only cases from the iPSYCH cohort, excluding all 2,304 comorbid cases (“ADHD vs ASD GWAS”). Using LDSC ^17^ we found an intercept of 0.9863 and a SNP-heritability of 0.4468 on the observed scale. Five genome-wide significant loci were identified, three of which have not previously been identified in GWAS of either of the two disorders separately, while all have been reported in related disorders and, remarkably, all but one are associated with cognitive abilities and/or neuroticism or neuroticism sub-items (**Table 2, Figure 1, Supplemental Table S5**). The lead variants all show opposite directions of effects in the two disorders.

**Table 2:**
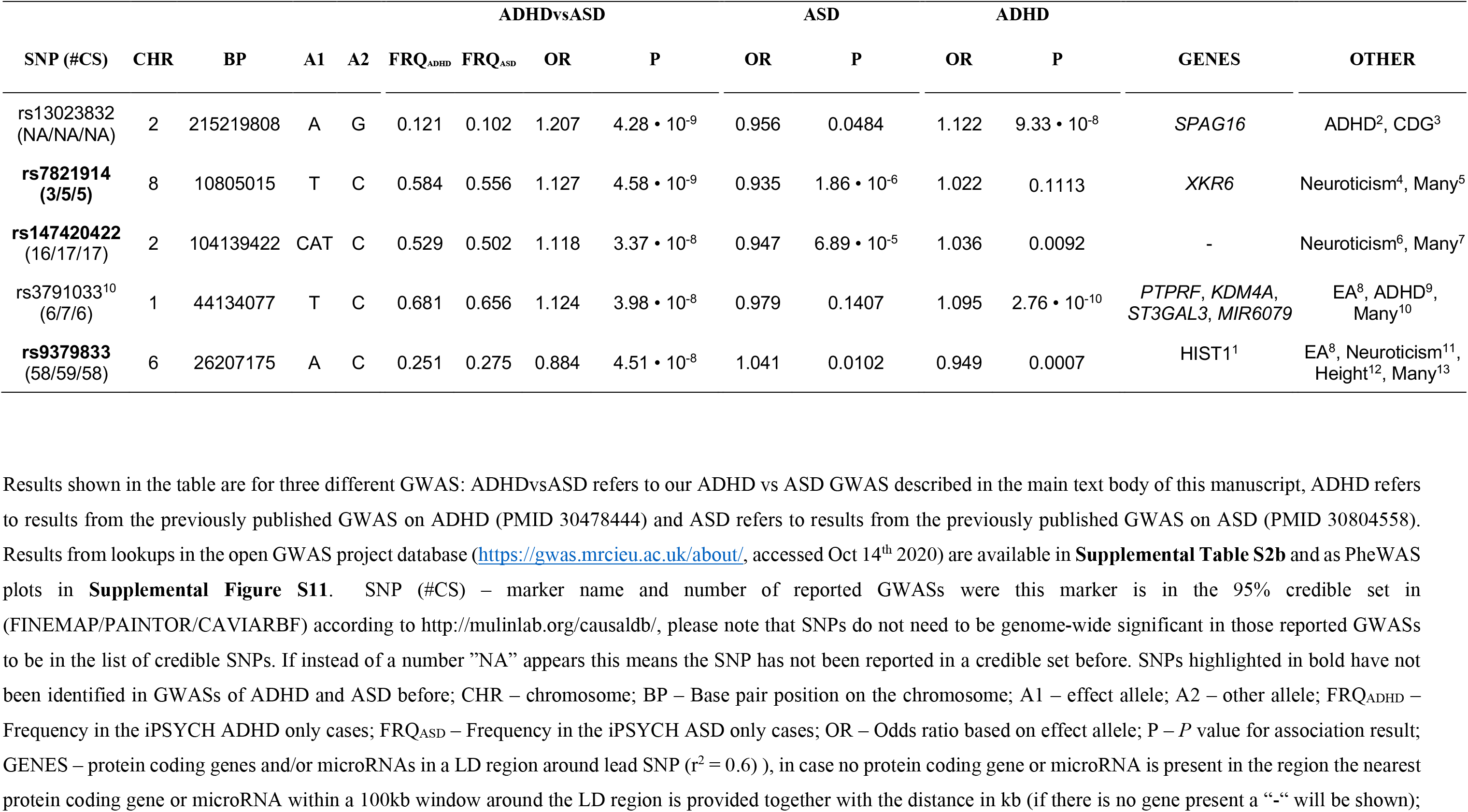
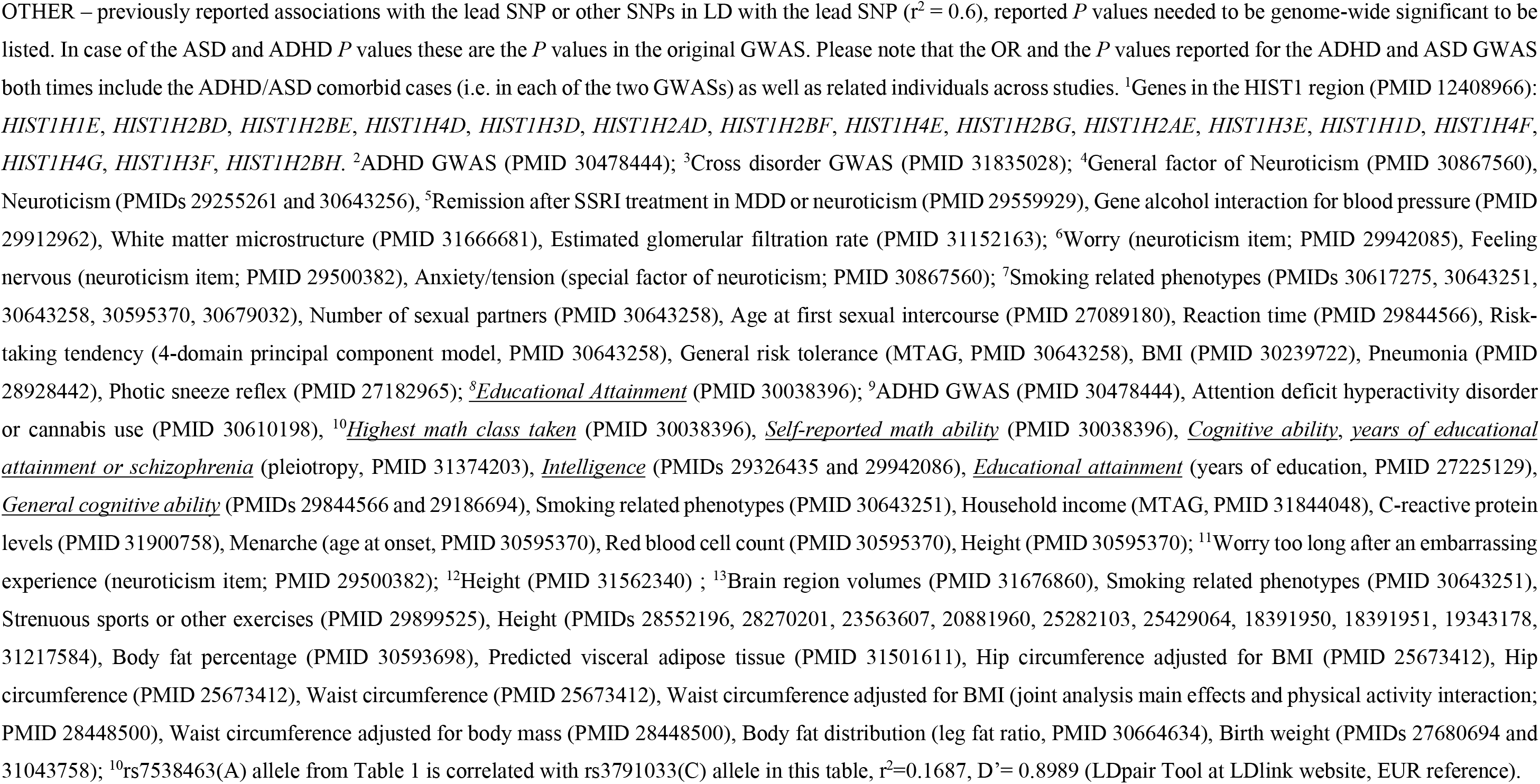
Results of differentiating GWAS (ADHD vs ASD)

Two of the five lead SNPs have previously been found associated with educational attainment ^24^. For the first SNP (rs3791033 on chromosome 1; *p* = 4.65×10^−23^) the C allele confers an increased risk for ASD and increased cognitive performance while the ADHD risk allele (T) is associated with decreased performance. Similarly, for the second SNP (rs9379833 on chromosome 6; *p* = 2.26×10^−8^) the A allele confers an increased risk for ASD and increased cognitive performance while the ADHD risk allele (C) is associated with decreased performance. Notably, this SNP (rs9379833) is located in the large histone gene cluster HIST1 ^25^ and has previously been associated with regional brain volume, specifically of the left globus pallidus ^26^ (*p* = 2.95×10^−8^; the C allele confers an increased risk for ADHD and a decreased volume while the ASD risk allele (A) is associated with an increased volume). Globus pallidus is part of the basal ganglia, which are involved in both motor and non-motor functions, including higher order cognition, social interactions, speech, repetitive behaviors and tics ^27^. It is also of note, that the lead SNP on chromosome 8 (rs7821914) is associated with neuroticism ^28^ (*p* = 9.46 ×10^−21^). For this SNP, the effect allele (C) in the neuroticism GWAS leads to an increased risk for ASD and a decreased risk for ADHD. An additional two of our lead SNPs are in LD (r^2^ > 0.6) with SNPs that have previously been identified in neuroticism or one of its subdimensions (rs147420422 and rs9379833; see **Table 2**). Results from additional lookups in the open GWAS project database (https://gwas.mrcieu.ac.uk/about/, accessed Oct 14^th^ 2020) are available in **Supplemental Table S5** and as PheWAS plots in **Supplemental Figure S6**.

TWAS using EpiXcan identified 11 Bonferroni significant genes/isoforms and 96 significant transcripts at FDR < 0.05 with different imputed expression in DLPFC between ADHD and ASD cases (**Supplemental Table S2** and **Figure 1**). The *HIST1H2BD-201* isoform located in the chromosome 6 (HIST1) GWAS locus showed the strongest association (*p* = 2.08×10^−9^) with higher expression in ADHD compared to ASD cases (**Supplemental Figure S3b**). The other genes/isoforms showed orders of magnitude less significant association, appointing *HIST1H2BD-201* as the top-ranking causal candidate in the locus. The remaining 10 Bonferroni significant genes/isoforms were located in the chromosome 8 GWAS locus (*SLC35G5-201, AF131215*.*5, AF131215*.*5-201, FAM167A* and *TDH-204*) or in two loci on chromosome 3 (3p21.1: *RFT1-204* and 3p21.31: *CAMKV-210, MON1A-201, RBM6-210* and *TRAIP*; **Supplemental Figures S3c + 3d**, respectively) where all except *TRAIP* were also genome-wide significant in gene-based analysis using MAGMA (**Supplemental Figure S4b** and **Supplemental Table S3**).

### Genetic correlations with other traits

To examine the polygenic architecture of the identified shared and differentiating genetic risk for the disorders we investigated the genetic correlations with 258 traits from a manually curated list of previously published GWAS and 597 traits from the UK Biobank making use of LD Hub ^29^ and LDSC ^30^. Among the 258 previously reported GWAS, 30 (combined GWAS) and 32 (ADHD vs ASD) traits showed significant correlations after Bonferroni correction for multiple testing (**Supplemental Table S4**). The strongest correlations for the liability differentiating ADHD vs ASD GWAS were observed for cognitive traits such as *years of schooling* (*r*_*G*_ = -0.669, *p*_corr_ = 3.68×10^−85^) and *childhood IQ* (*r*_*G*_ = -0.609, *p*_corr_ = 2.78×10^−10^), while the strongest correlations for the combined GWAS were with traits such as *depressive symptoms* (*r*_*G*_ = 0.506, *p*_corr_ = 2.08×10^−19^) and the cross-disorder analyses of the PGC (*r*_*G*_ = 0.433, *p*_corr_ = 5.30×10^−25^). Unsurprisingly, the largest difference in *r*_*G*_ (abs(Δ*r*_*G*_)) for the original ADHD and ASD GWAS was identified for a series of cognitive traits (largest abs(Δ*r*_*G*_) = 0.733 for *years of schooling*) (**Supplemental Table S4**). For a scatter plot of all genetic correlations that showed Z > 2 in the original ADHD and ASD GWAS please refer to **Supplemental Figure S7**.

### Tissue and cell-type enrichment analyses

We next tested whether genetic associations of shared and differentiating genetic underpinnings were enriched with respect to the transcriptomic profiles of human tissues. We found that the transcriptomic profiles related to brain tissues were significantly associated with the shared ADHD-ASD genetics (**Supplemental Figure S8**). At the general tissue level, these associations were with brain, pituitary and testis tissue. At the individual tissue level, the most significant association was observed for the basal ganglia (putamen), followed by the cerebellum. Cell-type enrichment analyses revealed experiment-wide significant association (across all data sets tested) of the red nucleus in the midbrain in human adult brain samples reported in La Manno et al. 2016 ^31^ (**Supplemental Figure S9c**). Associations that were significant within one of the three tested data sets individually, but not overall, were observed for several cell types, including e.g. dopaminergic and GABAergic neurons. For the disorder-differentiating analysis (ADHD vs ASD) we observed no significant association with tissues or specific cell-types after correction for multiple testing (**Supplemental Figure S10 and S11**). We also intersected our genetic associations with a recent multi-omic single-cell epigenetic catalogue of the human brain (obtained from GEO GSE147672 ^32^). Here both the combined and differentiating GWAS results showed significant enrichment for several neuronal cell populations (see **Supplemental Figure S12** and **Supplemental Table S6**), including excitatory and inhibitory neurons. Interestingly, the only difference in terms of significant associations between the combined and ADHD vs ASD GWAS was seen for oligodendrocytes (which were not significant in the combined but in the ADHD vs ASD GWAS). While aberrant myelination by oligodendrocytes resulting in disruption of white matter development has previously been reported in both ASD and ADHD ^33,34^, the degree of severity of this alteration might be a distinct pathophysiological factor ^35^.

### Polygenic characterization of case subgroups

We used two complementary polygenic risk score (PRS) approaches to investigate differences in polygenic load for ADHD, ASD and related phenotypes in the iPSYCH data across the three phenotypic subgroups: ASD-only, ADHD-only and comorbid cases. The multivariate PRS framework revealed significant association of the ASD-only subgroup with PRS for ASD (*p* = 6.89×10^−26^) and the ADHD-only subgroup with PRS for ADHD (*p* = 3.29×10^−23^). Both scores were trained with PGC-only GWAS results^5,36^. Strikingly, the ASD-PRS load on comorbid ASD+ADHD cases was similar to that on ASD-only cases (*p* = 0.77) and, likewise, the ADHD-PRS load on the comorbid subgroup was similar to that on ADHD-only cases (*p* = 0.44, **Figure 2)**, demonstrating that the comorbid cases carry a load of both ADHD and ASD polygenic scores that are similar to the load carried by the single-disorder cases of their respective disorder PRS. In other words, comorbid cases are double-burdened with both ASD and ADHD PRS. In contrast, the ASD-PRS load on ADHD-only cases was not different from controls (*p* = 0.79) and the ADHD-PRS was only slightly increased in ASD-only cases compared to controls (*p* = 3.26 × 10^−3^, **Figure 2**).

**Figure 2:**
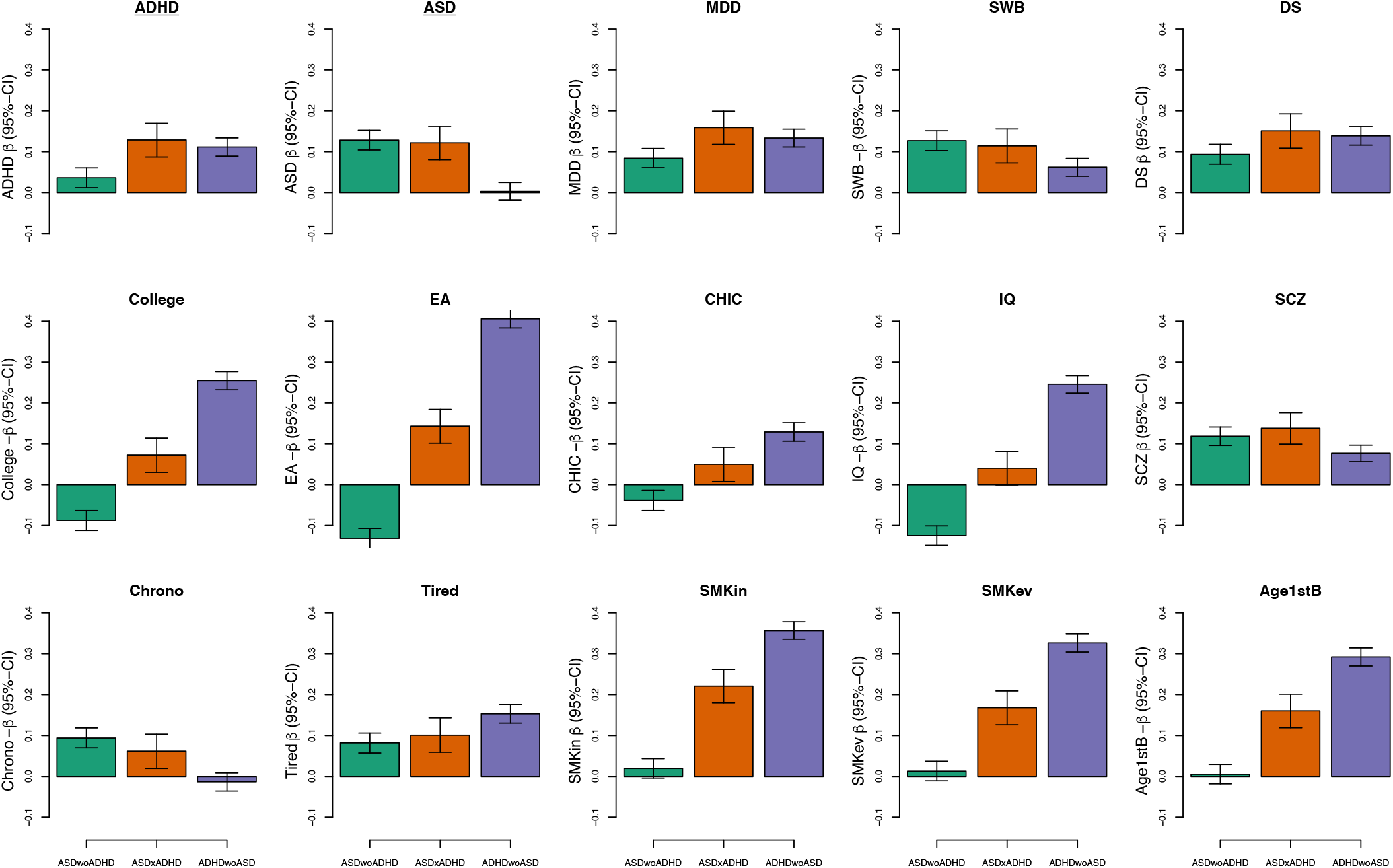
Multivariate PRS analyses for 15 traits associated with ADHD and/or ASD. Comparison of PRSs profiles across ADHD/ASD subtypes for 15 traits/phenotypes that have shown significant genetic correlation with ADHD and ASD in the past. Green bars represent ASD only cases, orange bars depict comorbid samples, and purple bars show average PRS for ADHD only cases. **<u>ADHD</u>** – attention-deficit/ hyperactivity disorder [PMID 20732625]; **<u>ASD</u>** – autism spectrum disorder [30804558 without the iPSYCH sample]; **MDD** – major depressive disorder [29700475 wo DK, wo 23am]; **SWB** – subjective well-being [27089181]; **DS** – depressive symptoms [27089181]; **College** – college completion [27046643]; **Edu** – educational attainment [30038396]; **CHIC** – childhood IQ [23358156]; **IQ** – IQ [29942086]; **SCZ** – schizophrenia [PGC3 woDK]; **Chrono** – chronotype [30696823]; **Tired** – self-reported tiredness [28194004]; **SMKos** – smoking initiation [30643251]; **SMKev** – ever smoker [30643258]; **Age1stB** – age of first birth [20418890].

Results from our leave-one-out framework analysis (including both PGC and iPSYCH data in the training GWAS) showed similar results (**Table 3**), adding further support to the observation of comorbid cases being double-burdened with both ASD and ADHD PRS. We note that in this analysis, the ASD-PRS load on ADHD-only cases as well as the ADHD-PRS load on ASD-only cases were increased compared to controls. Furthermore, secondary analysis in the leave-one-out framework suggested that ADHD cases with (*n* = 625) and without mild ID (*n* = 11,339) did not differ in terms of PRS for either ADHD or ASD. On the other hand, ASD cases with ID (*n* = 634) had lower PRS_ASD_ (OR = 0.89 [0.81-0.97], *p* = 0.0072) compared to those without mild ID (*n* = 8,681) but did not differ in terms of PRS_ADHD_ (**Table 3**).

**Table 3:** Results of ADHD and ASD polygenic risk score analyses in the iPSYCH cohort using a leave-one-out analysis framework.

| Cases<br>(coded as 1) | Comparison<br>(coded as 0) | PRS<sub>ADHD</sub> |  |  |  | PRS<sub>ASD</sub> |  |  |  |
| --- | --- | --- | --- | --- | --- | --- | --- | --- | --- |
|  |  | OR | LCI | UCI | P | OR | LCI | UCI | P |
| ADHD-only | Controls | 1.45 | 1.41 | 1.48 | $1.3 \cdot 10^{-207}$ | 1.08 | 1.06 | 1.11 | $7.5 \cdot 10^{-12}$ |
| ASD-only | Controls | 1.10 | 1.07 | 1.13 | $3.1 \cdot 10^{-13}$ | 1.21 | 1.18 | 1.24 | $1.2 \cdot 10^{-48}$ |
| Comorbid | Controls | 1.32 | 1.25 | 1.39 | $2.8 \cdot 10^{-25}$ | 1.22 | 1.16 | 1.29 | $3.5 \cdot 10^{-14}$ |
| Comorbid | ADHD-only | 0.92 | 0.88 | 0.97 | 0.0015 | 1.13 | 1.08 | 1.19 | $4.7 \cdot 10^{-7}$ |
| Comorbid | ASD-only | 1.22 | 1.16 | 1.28 | $6.4 \cdot 10^{-16}$ | 1.01 | 0.96 | 1.06 | 0.68 |
| ASD-only | ADHD-only | 0.76 | 0.74 | 0.78 | $4.5 \cdot 10^{-79}$ | 1.12 | 1.09 | 1.15 | $1.2 \cdot 10^{-15}$ |
| ADHD+ID | ADHD-no-ID | 0.97 | 0.88 | 1.06 | 0.46 | 0.94 | 0.86 | 1.03 | 0.19 |
| ASD+ID | ASD-no-ID | 1.03 | 0.93 | 1.12 | 0.58 | 0.89 | 0.81 | 0.97 | 0.0072 |

Results for per wave polygenic risk score analyses. PRS<sub>ADHD</sub> – Analyses using a polygenic risk score trained on an ADHD phenotype, PRS<sub>ASD</sub> – Analyses using a polygenic risk score trained on an ASD phenotype. Cases – group coded as 1 (cases) for the purpose of the analyses, Comparison – other group coded as 0 for the purpose of the analyses, OR – Odds ratio, LCI – lower boundary for 95% confidence interval, UCI – upper boundary for 95% confidence interval, P – P-value. Groups are as follows: ADHD-only – cases with ADHD diagnosis and without comorbid ASD diagnosis, ASD-only – cases with ASD diagnosis and without comorbid ADHD diagnosis, Comorbid – cases with comorbid ADHD and ASD diagnoses, Controls – individuals without ADHD and ASD diagnoses. P-values shown are without correction for multiple testing. Experiment-wide significant at 0.0042 (Bonferroni corrected for 2 x 6 tests). Additional secondary analyses also compare groups of individuals with ADHD or ASD with co-occurring mild intellectual disability (ADHD+ID and ASD+ID) to those without (ADHD-no-ID and ASD-no-ID).

To further dissect the genetic architecture across the ASD and ADHD subgroups we examined the relative burden of PRS for phenotypes and traits that have shown significant genetic correlation with ADHD and ASD ^5,6 37^ using the multivariate framework analysis. While PRS for SZ and depression (and genetically related phenotypes) did not show substantially different loads across the subgroups, other traits showed compelling differences (**Figure 2**). For instance, years of education, IQ, age at first birth, tiredness, and smoking showed differences between ADHD-only and ASD-only cases with the comorbid cases at an intermediate level. These differences were most compelling for the cognitive phenotypes displaying PRS loads in opposite directions for the single-disorder cases and intermediate loads for comorbid cases. In addition, analyses for the chronotype trait showed similar PRS loads in ASD-only and comorbid cases without evidence for loading in ADHD-only cases, reflecting that chronotype is genetically correlated to ASD but not ADHD ^5,6^.

An item-level analysis of neuroticism revealed specific patterns of associations for the ASD-only and ADHD-only groups that were mostly consistent with previously described patterns ^37^ (**Supplemental Figure S13**). On average, ADHD-only cases showed much stronger association than ASD-only cases with items belonging to the depressed affect cluster (e.g. the *MOOD* item) compared to the worry cluster (for a definition of the clusters see **Supplemental Figure S13**). For comorbid cases a distinct pattern was observed with PRS loads either ranking between the ADHD- and ASD-only cases (e.g. for the *MOOD* item) or even exceeding the two single-disorder groups (e.g. for the *GUILT* item).

Summarizing, we observe a genetic architecture of comorbid cases that presents itself in clear distinction from the ADHD and ASD single-disorder cases. Showing burden of both ASD and ADHD genetic risk, the comorbid cases also carry polygenic load profiles across other phenotypes that distinguishes them from their single-disorder cases, typically by carrying an intermediate load level but in some cases a load similar to just one of the single-disorder groups.

### Genetic correlation and heritability across case subgroups

We recently reported an LDSC genetic correlation of 0.36 between ASD and ADHD using the largest GWAS meta-analyses of the two disorders, including multiple cohorts and comorbid cases ^5^. Here we investigated the correlations across diagnostic subgroups of the disorders in the iPSYCH sample using the GREML approach of GCTA ^38^. For ASD and ADHD overall, we found *r*_*G*_ = 0.497 (SE = 0.054, *p* = 7.8×10^−19^). Excluding the comorbid cases reduced the correlation to *r*_*G*_ = 0.397 (SE = 0.056, *p* = 6.3×10^−12^). After excluding cases with intellectual disability (ID), the correlations between ASD and ADHD were even stronger: *r*_*G*_ = 0.523 (SE = 0.054, *p* = 6.5×10^−21^) and *r*_*G*_ = 0.425 (SE = 0.056, *p* = 1.7×10^−13^) with and without comorbid cases, respectively. All the GCTA results on genetic correlations and SNP heritability estimates can be found in **Supplemental Table S7** and **Supplemental Figure S14**.

Correlations between ADHD and ICD-10 diagnostic subcategories of childhood autism (F84.0, *n* = 3,273), atypical autism (F84.1, *n* = 1,472), Asperger’s syndrome (F84.5, *n* = 4,363), and other/unspecified pervasive developmental disorders (other PDDs, F84.8-9, *n* = 3,794), reducing to nonoverlapping groups when performing pairwise comparisons (**Supplemental Table S7**), were mostly similar to those for the ASD group overall, albeit with generally higher estimates for the groups with other PDDs and Asperger’s syndrome (**Supplemental Table S7** and **Supplemental Figure S14**).

## Discussion

This study dissects the genetic architecture for shared and differentiating genetic underpinnings of ADHD and ASD as well as across case subgroups. At the single variant level, we identified novel shared loci for the two disorders and, to the best of our knowledge, the first genome-wide significant loci differentiating the disorders. Integration with DLPFC transcriptomic data identified and prioritized several possibly causal genes (**Box 1**). At the polygenic level, we revealed compelling differences across comorbid and single-disorder case groups.

### Box 1

Prioritized genes or transcripts that are (i) located in GWAS loci and/or are genome-wide significant in gene-wise analysis and (ii) Bonferroni significant in TWAS. Genes/transcripts showing increased imputed DLPFC expression in ADHD-ASD combined compared to controls are highlighted in red while those with decreased expression are blue. Genes/transcripts showing decreased imputed DLPFC expression in ASD compared to ADHD are highlighted in green while those with decreased expression in ADHD compared to ASD are purple.

#### Shared liability genes identified in the combined ADHD-ASD GWAS and TWAS

*Keratin 8 Pseudogene 46 (KRT8P46) (transcript, isoform 201)*: Located on chromosome 4 (**Supplemental Figure 3a**), *KRT8P46* is a pseudogene located in an intron of *MANBA*, a gene that has been previously associated with ADHD and asthma ^1^ as well as blood protein levels ^2^. Pseudogenes have recently been highlighted as regulators in health and disease ^3,4^ amongst others through potential regulatory relationship with their parent genes. It is of note that another keratin 8 pseudogene (*KRT8P44*) is located in a region that has been identified to harbor a rare CNV associated with ADHD^5^.

#### Differentiating liability genes identified in the ADHD vs ASD GWAS and TWAS

*HIST1H2BD (transcript, isoform 201)*: Located in the cytogenetic band 6p22.2 as part of the histone gene cluster (more precisely the H1 histone family) (**Supplemental Figure 3b**). The gene (also known as *H2BC5*) encodes the Histone H2B type 1-D protein. Histone proteins in general are involved in the structure of chromatin in eukaryotic cells and play a central role in transcription regulation, DNA repair, DNA replication, and chromosomal stability. Little is known about the specific function of *HIST1H2BD*, however, deleterious de novo mutations in several histone modifying or interacting genes (albeit not including *HIST1H2BD)* ^*6-8*^ as well as in core histone genes ^7,9^ have been associated with autism and developmental delay with autistic features.

*CAMKV (transcript, isoform 210)*: Located together with two other TWAS significant genes in a region on chromosome 3p21.31 (**Supplemental Figure 3c**), which has been reported to harbor CNVs in individuals with autism, intellectual disability and developmental delays ^10-13^. The calmodulin kinase-like vesicle-associated (CaMKv) is a pseudokinase required for the activity-dependent maintenance of dendritic spines ^14^.

*RFT1(transcript, isoform 204)*: Located on chromosome 3p21.1 (**Supplemental Figure 3d**) *RTF1* encodes an enzyme which catalyzes the translocation of the Man(5)GlcNAc (2)-PP-Dol intermediate from the cytoplasmic to the luminal side of the endoplasmic reticulum membrane in the pathway for the N-glycosylation of proteins. Mutations in *RFT1* cause recessive congenital disorder of glycosylation type 1N (CDG1N) <u>[MIM:612015]</u>, which presents with a wide variety of clinical features, including severe developmental delay, hypotonia, dysmorphic features and epilepsy.

*FAM167A (transcript, isoform 201)*: Located together with three other TWAS significant genes in the identified GWAS locus on chromosome 8 (**Supplemental Figure 3e**). Using data from a subset of our iPSYCH ASD samples, a previous study identified two differentially methylated positions (DMPs) in the same region that showed association with polygenic risk for ASD ^15^. One of the DMPs was annotated to the Family With Sequence Similarity 167 Member A gene (*FAM167A*), while the other was annotated to *RP1L1*.

The identified shared loci are generally highly pleiotropic and have previously been identified in GWAS of related disorders or cross-disorder studies including ADHD and/or ASD. However, considering only psychiatric disorders, the two novel shared loci (on chromosome 1 and 4) appear to be shared only between ADHD and ASD (**Table 1**). This is consistent with evidence from structural equational modeling of eight major psychiatric disorders, showing that ASD and ADHD cluster together in a group of early-onset neurodevelopmental disorders along with Tourette syndrome ^8^.

Analyzing genetic correlation with other traits, the combined GWAS results showed strong correlations of shared ADHD-ASD genetics with other psychiatric phenotypes, suggesting that additional shared loci that may be discovered with increasing sample sizes in future studies will likely show a high degree of overlap with other psychiatric disorders, similarly to the shared loci reported here.

In the ADHD vs ASD GWAS we identified five genome-wide significant loci, all showing opposite allelic directions in the separate GWAS of the two disorders, providing the first specific genetic clues to understanding the biology that drives the pathophysiology towards developing one or the other disorder. The top-ranking TWAS gene/isoform was *HIST1H2BD-201*, which was two orders of magnitude more significant than the second-ranking (*CAMKV-210*) and the only Bonferroni significant transcript in the identified histone 1 GWAS locus. Deleterious *de novo* mutations in several histone modifying or interacting genes ^39-41^ as well as in core histone genes ^40,42^ have been associated with autism and developmental delay with autistic features. The haploinsufficiency resulting from these de novo mutations is consistent with our TWAS result showing reduced expression of *HIST1H2BD-201* in ASD (relative to ADHD). Intriguingly, the ASD risk allele of the lead SNP in the locus is also associated with both increased educational performance ^24^ and increased volume of the left globus pallidus ^26^ while the opposite is the case for the ADHD risk allele. As part of the basal ganglia, globus pallidus is involved in several functions relating to phenotypic domains affected in ASD and/or ADHD such as cognition, social interactions, speech, repetitive behaviors and tics ^27^. Taken together our results suggest that the identified ADHD-ASD differentiating locus on chromosome 6 has downstream effects involving differential expression of the histone isoform *HIST1H2BD-201* and volumetric changes of the left globus pallidus in driving the pathophysiology towards either ASD or ADHD and impacting key phenotypic domains such as educational performance, social interaction and motor impairments.

Previous studies found ASD and ADHD to display opposite genetic correlations with cognitive traits like educational attainment when assessing common variants genome-wide ^5,6,43^. Corroborating these reports, we found that the ADHD vs ASD GWAS showed the strongest correlations for cognitive traits among the multiple phenotypes tested (**Supplemental Table S4 and Supplemental Figure S7**). Moreover, two of the identified differentiating loci (on chromosome 1 and 6) have lead SNPs that are genome-wide significant in educational attainment and show opposite allelic effects with increasing and decreasing educational performance for the ASD and ADHD risk alleles, respectively.

We note that the chromosome 1 locus (at position 44Mb) was identified, counterintuitively, in both the shared and differentiating GWAS albeit with different lead SNPs (**Table 1 and 2**). The locus covers a gene-rich 250kb region of generally strong linkage disequilibrium (LD) but it also harbors variants with limited LD to the main haploblock (**Supplemental Figure S1a and S5d**). The two lead SNPs are located 62kb apart and show low pairwise LD (*r*^*2*^ = 0.1687, **Table 2**), indicating that the two SNPs are largely independent markers for association. This LD difference is also reflected in the different lists of other traits with previously reported associations for the lead SNPs or their LD proxies (**Table 1 and 2**). Furthermore, the locus was the single locus showing significant heterogeneity across cohorts in the recent ADHD GWAS ^6^ where the 23andMe sample provided no support for the otherwise consistently supported locus and, also in contrast to the other cohorts, exhibited limited genetic correlation with educational attainment.

Our analyses revealed the expected enrichment of brain-expressed genes for the combined GWAS. In particular, the basal ganglia and the cerebellum seem to be implicated. Both structures have been found to be altered in both ASD ^27,44^ and ADHD ^45-47^, with evidence for reductions in basal ganglia volume the most robustly observed finding in the neuroimaging literature for both ASD and ADHD. In addition to the brain, enrichment of genes expressed in the pituitary gland and the testes was also observed for the combined GWAS results. This finding may suggest the involvement of the (hypothalamic-)pituitary-gonadal axis and potentially estrogen signaling, which is known to play a role in psychiatric disorders, cognition, and neuroprotection via several neurotransmitter systems, such as the dopaminergic, serotonergic, and glutamatergic system (e.g. Hwang et al. ^48^). The cell-type enrichment result implicating the red nucleus in midbrain is also consistent with our knowledge of phenotypic sharing between ASD and ADHD, as it relates to skilled movements and motor control in the limbs as well as jaw: both motor coordination problems and speech problems are frequent in both ASD and ADHD ^49,50^. The red nucleus is strongly connected with many brain structures involved in ASD and ADHD, including the basal ganglia and the cerebellum ^51^.

Dissecting the polygenic architecture using PRS approaches we observed remarkable differences across comorbid and single-disorder case groups. The comorbid cases carry a double burden of ASD- and ADHD-PRS, whereas the single-disorder cases were largely just (single-)burdened for the respective disorder. Thus, cases diagnosed with both disorders have on average a similar level of genetic liability to each disorder as the single-disorder cases, providing strong biological support for the change in diagnostic guidelines from DSM-IV to DSM-5 allowing for diagnoses of both disorders in the same person. It also supports pharmacological treatment of comorbid ADHD in individuals with ASD. In a recent meta-analysis, 25-32% of individuals with ASD also fulfill criteria for ADHD ^13^, yet only 15-16% are treated with ADHD medications ^52,53^, despite strong evidence of beneficial effects on the core symptoms of ADHD and potentially also reduced risk of injuries ^54^, depression ^55^, suicidal behavior ^56^ and improved academic performance ^57^. Moreover, it indicates that pharmacological treatment of symptoms like hyperactivity, inattention, impulsivity, aggression and tics in cases diagnosed with either ADHD or ASD may be guided by the individual symptomatology regardless of the given diagnosis.

The multivariate PRS analysis also revealed clear differences across the case subgroups for PRS from several other traits, particularly cognition-related traits, again highlighting the opposite relationship for the two disorders with e.g. educational attainment and IQ, which, unsurprisingly, was expanded for the single-disorder cases (with exclusion of the comorbid cases), while the PRS load in the comorbid case group was placed in-between but dominated by the strong negative correlation observed for ADHD cases.

We recently reported a significant genetic correlation of *r*_*G*_ = 0.36 between ASD and ADHD, using LDSC and results from GWASs that included multiple cohorts and comorbid cases ^5^. This was a considerable increase from the previous estimate of *r*_*G*_ = 0.08 (SE = 0.10, *p* = 0.40), which was based on much smaller GWAS sample sizes without information on comorbid diagnoses ^58^. Here we analyzed exclusively the iPSYCH cohort, which is relatively homogeneous and has information on all diagnoses given to each individual. We found a higher correlation (*r*_*G*_ = 0.497), which remained substantial when excluding the comorbid cases (*r*_*G*_ = 0.397), demonstrating that the genetic overlap between the disorders is far from driven by comorbid cases alone. This is corroborated by data from Swedish twin studies that supports the distinction of ASD and ADHD, but also suggests considerable co-occurrence of symptoms of both disorders in individuals only fulfilling diagnostic criteria for one of the two disorders ^59,60^.

In addition, the correlations increased when excluding cases with ID, indicating that cases with ID are more genetic heterogeneous in common variant risk between the two disorders than cases without ID. A recent exome sequencing study of ASD and ADHD (also in the iPSYCH cohort) showed that the disorders have substantial overlap in rare variant risk and that cases with ID carry a higher load of (ultra)rare damaging risk variants compared to cases without ID ^10^. Consistent with this, our PRS analyses found that there was lower ASD PRS in the group of ASD cases with comorbid mild ID (IQ=50-70) compared to those without mild ID. Taken together, these observations are consistent with the notion that the genetics differentiating the two disorders may be driven primarily by common variants and more extensively for cases with ID than without ID. However, larger sample sizes for both GWAS and sequencing studies are needed to clarify this.

In conclusion, we have disentangled the shared and differentiating genetic liability underlying ASD and ADHD, identifying novel shared as well as disorder-specific risk variants informing on the pathophysiology. In addition, we have revealed specific patterns of polygenic architecture that are characteristic for comorbid cases compared to single-disorder cases. The results advance the understanding of the complex etiologic basis and relationship between ASD and ADHD towards the long term goals of better diagnosis and treatment of these disorders.

## Methods

We report results from a framework of different analyses all carried out in large-scale samples from the Psychiatric Genomics Consortium (PGC) and the Lundbeck initiative of integrative psychiatric research (iPSYCH) samples. We used samples included in the most recent GWAS of ASD ^5^ and ADHD ^6^. For the purpose of this manuscript we will refer to individuals in the study cohort (most importantly in the iPSYCH cohort) that at the time of inclusion only had one of the two diagnoses registered (i.e. ADHD or ASD) as *ADHD-only* and *ASD-only* cases, respectively. We refer to individuals that during their lifetime and up to the time of inclusion had both an ADHD and ASD diagnosis registered as *comorbid* cases. Furthermore, we refer to these three groups of cases (i.e. *ADHD-only, ASD-only*, and *comorbid*) as *ASD and ADHD subgroups*.

### Sample description and additional quality control

Details about study specific case and control selection criteria and how individuals were drawn from the overall population-based iPSYCH case-cohort sample ^61^ can be found in the respective publications ^5,6^. Here we focus on important differences in the case and control selection criteria in the iPSYCH cohort as well as additional quality control (QC) procedures necessary for the current study.

Almost all of the inclusion and exclusion criteria for the original studies were also used in this study. The only difference compared to the original studies was an additional exclusion criterion that removed individuals with a moderate to severe mental retardation (ICD10: F71-F79) from both the case and control cohorts. While this criterion was also used in the original ADHD GWAS ^6^, it was, however, not used in the original ASD GWAS ^5^. The rationale for this decision lies in the interpretability of our results where we treated ADHD and ASD consistently. We address the potential impact of this decision through different analyses (see **Table 3, Supplementary Figure S14b**, and **Supplementary Table S7**).

Wave-wise pre-imputation QC and imputation of the iPSYCH case-cohort sample were taken from the original ADHD and ASD GWAS, respectively. Details about the respective steps and filters can be found elsewhere ^5,6^. Since our analysis framework used a combined study cohort with samples from both the original ADHD and ASD GWAS we performed some additional QC on the combined sample. The additional QC steps included the removal of related individuals across the original ADHD and ASD GWAS as well as a new principal component analysis (PCA) on the combined sample after exclusion of these related individuals. Following the same procedures as in the original studies, pairs of subjects were identified with pi-hat> 0.2 (using PLINK’s ^62^ identity by state analysis) and one subject of each such pair was excluded at random (with a preference for keeping cases). PCA was carried out using smartPCA in the EIGENSOFT software package ^63,64^ using the framework of the Ricopili pipeline ^65^. The original PGC datasets for ADHD and ASD did not include overlapping individuals and therefore the original datasets and summary statistics were used. The final combined dataset across all samples comprised 34,462 cases (i.e. individuals with an ADHD and/or ASD diagnosis) and 41,201 controls. We only included samples of European ancestry from the original ADHD and ASD GWAS. Among the cases in the iPSYCH cohort 11,964 had an ADHD-only diagnosis, 9,315 had an ASD-only diagnosis, and 2,304 individuals had a comorbid diagnosis.

### Genome-wide association analyses

Like with the original GWAS in ADHD and ASD, all processing and analyses for the individual GWAS and meta-analyses (see below) used the framework of the Ricopili pipeline ^65^. More details on individual modules and steps can be found elsewhere ^5,6,65^. We ran two main GWAS within our framework of analyses. The first one aimed to identify shared genetic risk for ADHD and ASD (*combined GWAS*) and the second one aimed to identify differentiating genetic risk with an opposite direction of effects for ADHD and ASD (*ADHD vs ASD GWAS*). All analyses of the iPSYCH sample and meta-analyses with the PGC samples were conducted at the secured national GenomeDK high-performance computing cluster in Denmark. The study was approved by the Regional Scientific Ethics Committee in Denmark and the Danish Data Protection Agency.

### Combined GWAS

We first ran an analysis in the combined dataset, i.e. on all 34,462 cases and 41,201 controls. The GWAS was conducted in each cohort (i.e. in the wave-wise iPSYCH samples and the individual PGC cohorts) using logistic regression with the imputed additive genotype dosages. The first 5 principal components (PCs) were included as covariates to correct for population stratification (**Supplementary Information**), and variants with imputation INFO score < 0.8 or minor allele frequency (MAF) < 0.01 were excluded. The resulting summary statistic files were then meta-analyzed using an inverse-variance weighted fixed effects model ^66^. Post-processing of the summary statistics files through the Ricopili pipeline ^65^ created Manhattan plots, individual regional associations plots, and forest plots. For a QQ-plot of the analysis please refer to **Supplemental Figure S15a**.

### ADHD vs ASD GWAS

To identify unique genetic risk loci or loci with opposite direction of effects for ADHD and ASD we ran a case-only analysis for the ADHD-only (coded as 1, i.e. “pseudo-cases”; *n* = 11,964) against ASD-only cases (coded as 2, i.e. “pseudo-controls”; *n* = 9,315) in the iPSYCH cohort. We excluded the comorbid cases from this GWAS. Similar to the analysis in the combined sample (see above) GWAS was conducted wave-wise using logistic regression with the imputed additive genotype dosages. The first 5 PCs were included as covariates to correct for population stratification, and variants with imputation INFO score < 0.8 or MAF < 0.01 were excluded. The resulting summary statistic files were then meta-analyzed using an inverse-variance weighted fixed effects model ^66^ and visualization of results was achieved through the Ricopili pipeline ^65^. Post-processing of the summary statistics files through the Ricopili pipeline ^65^ created Manhattan plots, individual regional associations plots, and forest plots. For a QQ-plot of the analysis please refer to **Supplemental Figure S15b**.

### Identification of previously reported associations for top findings

Different resources were used to look up previously reported associations of our top findings with other phenotypes and traits within and outside of psychiatry. We assessed associations reported in the open GWAS project database (https://gwas.mrcieu.ac.uk/about/, accessed Oct 14^th^ 2020; see **Supplementary Table S1** and **Supplementary Table S5** for results) and also used the GWAS ATLAS website ^67^ to visualize PheWAS analyses (see **Supplementary Figures S2** and **S11**). Finally, we also used results from the GWAS Catalog ^68^ (see **Table 2**).

### Transcriptomic imputation model construction and transcriptome-wide association study (TWAS)

Transcriptomic imputation models were constructed as previously described ^20^ for dorso-lateral prefrontal cortex (DLPFC) transcript levels ^69^. The genetic dataset of the PsychENCODE cohort was uniformly processed for quality control (QC) steps before genotype imputation. We restricted our analysis to samples of European ancestry as previously described ^20^. Genotypes were imputed using the University of Michigan server ^70^ with the Haplotype Reference Consortium (HRC) reference panel ^71^. Gene expression information (both at the level of gene and transcript) was derived from RNA-seq counts which are adjusted for known and hidden confounds, followed by quantile normalization ^69^. For the construction of the transcriptomic imputation models we used EpiXcan ^20^, an elastic net based method, which weighs SNPs based on available epigenetic annotation information ^72^. EpiXcan was recently shown to increase power to identify genes under a causality model when compared to TWAS approaches that don’t integrate epigenetic information ^73^. We use this model (924 samples from DLPFC) due to power considerations ^20^; in comparison, brain gene expression imputation models based on GTEx V8 ^74^ are trained in 205 or fewer samples. We performed the transcript-trait association analysis for the traits in this study as previously described ^20^. Briefly, we applied the S-PrediXcan method ^20^ to integrate the GWAS summary statistics and the transcriptomic imputation models constructed above to obtain association results at both the level of genes and transcripts.

### Cell-type enrichment analysis

A major portion of cell type specific enrichment is attributed to distal regulatory elements, as local regulatory events remain highly consistent across various tissues and cell types ^75^. Therefore, we examined an overlap of common genetic variants of investigated traits (see **Supplemental Figure S14** and **Supplemental Table S6**) and open chromatin from scATAC-seq study (single-cell assay for transposase accessible chromatin) ^32^ using the LD-score partitioned heritability approach ^76^. All regions of open chromatin were extended by 500 base pairs in either direction. The broad MHC-region (hg19 chr6:25-35MB) was excluded due to its extensive and complex LD structure, but otherwise default parameters were used for the algorithm.

### Additional functional characterization and annotation of main findings

We used a number of different approaches combining in house scripts and data with those available via the FUMA v1.3.6a ^23^ website (http://fuma.ctglab.nl) for downstream functional characterization and annotation of our findings. For FUMA we uploaded our summary statistics from the individual analyses. We also used FUMA to perform tissue expression analyses on data available through their website. Finally, we used FUMA to perform cell-type specificity analyses ^77^ based on our summary statistics. For all above mentioned analyses default settings were applied. More detailed information about the individual third-party datasets (available through FUMA) included in the analyses as well as individual aspects of the FUMA analyses can be found in the **Supplemental information. Supplemental Table S8** contains results from standard FUMA-based analyses, such as eQTL and chromatin interaction mapping.

### Gene-based analysis

We also used FUMA v1.3.6a ^23^ to perform gene-based analysis. Genome-wide significance was assessed through Bonferroni correction for the number of genes tested. More detailed information about the individual third-party datasets (available through FUMA) included in the analyses as well as individual aspects of the gene-based analyses can be found in the **Supplemental information**.

### Our results in context of other findings

Since the publication of the original ADHD and ASD results a few studies have investigated the shared and unique risk architecture of these disorders. We compared our results with the findings of the cross disorder working group of the PGC ^8^ and a recent analysis based on structural equation modelling of 11 major psychiatric disorders [https://doi.org/10.1101/2020.09.22.20196089]. We also compared our results with recent analyses that aimed at identifying disorder-specific SNPs for psychiatric disorders ^78,79^.

### Polygenic risk score (PRS) analyses

To examine potential polygenic heterogeneity across ADHD and ASD subtypes, we investigated how PRS trained on different phenotypes were distributed across ADHD-only, ASD-only and comorbid subgroups in the iPSYCH data through two complementary analysis frameworks: multivariate PRS and leave-one-out PRS. These two approaches have different strengths and limitations, allowing for robust interrogation of differences in ADHD and ASD subgroups in terms of polygenic burden for ADHD and ASD, as well as genetically related phenotypes.

### Multivariate PRS analyses

To examine the relative burden of PRS for phenotypes and traits that have shown significant genetic correlation with ADHD and ASD in the past ^5,6,37^ across ADHD and ASD subgroups in the iPSYCH data, we ran a multivariate regression of the scores on these subgroups, adjusting for PCs and batch. For details, see Grove et al. ^5^. In brief, this is a regression of multiple outcome variables and can superficially be viewed as running a linear regression for each score on the ADHD and ASD subgroups. It allows us to compare the average PRS across subtypes for scores from a number of phenotypes while accounting for the inherent correlation between scores and adjusting for necessary covariates. This enables testing a whole array of hypotheses comparing both subtypes and PRS. Polygenic scores were generated by clumping and thresholding employing standard Ricopili settings as explained in ^5^ and using summary statistics from the GWASs ^5,36,80-89^.

### Leave-one-out PRS analyses

As a complementary approach, a leave-one-wave-out approach within the iPSYCH data was used to maximize power and maintain independent target and discovery samples for PRS analyses. Meta-analyses were run in METAL (using inverse-variance weighted fixed effects models with the STDERR scheme), including the per-wave GWAS summary results from all but one wave of data, for each combination of waves. Separate meta-analyses were run for GWAS of ADHD-only (excluding comorbid ASD or severe ID, defined as IQ ≤ 50) cases vs. controls and ASD-only (excluding comorbid ADHD or severe ID) cases vs. controls, using independent (split) controls. For each set of discovery results, LD-clumping was run in PLINK v.1.9 ^90^ (with the parameters --clump-kb 500 --clump-r2 0.3) to obtain a relatively independent set of SNPs, while retaining the most significant SNP in each LD block. The SNP selection p-value threshold used was *p* < 0.5. Asymmetric/ambiguous SNPs (AT, TA, CG, GC), indels, multi-allelic and duplicate position SNPs were excluded. SNPs with MAF < 0.01, INFO < 0.8 or present in less than half of the sample were filtered out. PRS for ADHD and ASD were calculated for each individual in each target wave by scoring the number of effect alleles weighted by the log(odds ratio [OR]) across the set of independent clumped, meta-analyzed SNPs in PLINK. PRS were derived in best guess imputed data after filtering out SNPs with MAF < 0.05 and INFO < 0.8. The PRS were standardized using z-score transformations; ORs can be interpreted as the increase in risk of the outcome, per standard deviation in PRS. Logistic regression analyses including 5 PCs were run to test for association of PRS with each of the outcomes within each wave, as follows: a) ADHD-only cases vs. controls, b) ASD-only cases vs. controls, c) comorbid cases vs. controls, d) ADHD-only cases vs. ASD-only cases, e) ADHD-only cases vs. comorbid cases, and f) ASD-only cases vs. comorbid cases. Cases were coded as 1 and controls as 0, except that comorbid cases were coded as 1 in case-case comparisons and in analysis (d), the ASD-only cases were coded as 1. Overall meta-analyses of these per-wave analyses were performed in R using the ‘metafor’ package. As secondary tests, we stratified the ADHD-only and ASD-only cases by presence of mild ID (defined as IQ between 50-70). We also examined differences across several ASD hierarchical subtypes (childhood autism, atypical autism, Asperger’s, and pervasive developmental disorders mixed; see Grove et al ^5^ and **Supplemental Table S7**). Several sensitivity tests were also run (including sex as a covariate, excluding cases and controls with mild ID).

### Genetic correlations (LDhub)

The genetic correlations of our different datasets with other phenotypes were evaluated using LD Score regression (LDSC) ^30^ and the LD Hub ^29^ website (http://ldsc.broadinstitute.org/ldhub/). In brief, we re-reran analyses of the original GWAS of ADHD and ASD ^5,6^ in the European-only datasets since new phenotypes have been added to LD Hub after publication of the original analyses. We also uploaded summary statistics for the two analyses described above, i.e. the GWAS in the combined sample (combined GWAS) and the pseudo case-control analysis (ADHD vs ASD GWAS). We used all available phenotypes in LD Hub ^29^ but performed analyses for the UKBB traits (*n* = 597) and the remaining individual phenotypes (*n* = 257) separately. For ADHD ^6^ and ASD ^5^ the most recent summary statistics replaced corresponding summary statistics in LD Hub as these had not been included at the date of analysis. The same was true for the summary statistics of major depressive disorder (MDD ^85^) and bipolar disorder (BD ^91^). Levels of experiment-wide significance (Bonferroni correction for number of tests applied) were also established separately within the two groups, i.e. in the UKBB traits (*p* < 8.38 x 10-5) and the remaining individual phenotypes (*p* < 0.00019), respectively.

### GCTA-GREML analyses across subgroups

The additive variance explained by our GWAS dataset (SNP-based heritability; SNP-h^2^) was estimated in the iPSYCH sample using the GREML approach of GCTA ^38^ for ADHD versus ASD and for ADHD versus each of the ASD sub-phenotypes listed in **Table 1**. The genetic relationship matrix (GRM) between all pairwise combinations of individuals was estimated using all case-control samples. The strict best-guess-genotypes (i.e. SNPs with INFO > 0.8, missing rate < 0.01 and MAF > 0.05, INDELs removed) were used for GRM estimation. GCTA-GREML accounts for linkage disequilibrium (LD) ^92^, and the GRM estimation was therefore performed on a non-LD-pruned dataset. Estimation of the phenotypic variance explained by the SNPs was performed for each of the phenotypes listed in **Supplemental Table S7**, with PCs 1-20 included as continuous covariates and wave (1-23) as categorical dummy variables. ADHD prevalence of 0.05 and ASD prevalence of 0.01 was assumed to estimate the variance explained on the liability scale. Prevalence was estimated for hierarchical ASD phenotypes based on the estimate for the overall ASD phenotype and the proportion of each hierarchical phenotype over all ASD cases observed in our sample. Genetic covariance between pairs of traits (**Supplemental Table S7**) was estimated using the bivariate approach implemented in GCTA, by randomly splitting controls into two groups, one for each trait, in proportions corresponding to the proportion of the cases for each of the two traits in the total sample. PCs 1-20 and dummy variables for wave 1-23 were included as covariates in the bivariate analyses. Two-tailed p-values were obtained for *r*_*G*_ point estimates based on the standard error estimated by GCTA using the approach by Altman and Bland ^93^.

GCTA-GREML analyses were conducted for ADHD versus ASD main diagnosis (**Supplemental Figure S5a**), by (1) excluding individuals with both phenotypes (comorbid) and (2) by randomly splitting comorbid cases into either ADHD or ASD. GCTA analyses were, in addition, conducted for ADHD versus four ASD sub-phenotypes, by (1) excluding individuals with both phenotypes (comorbid) and (2) by randomly splitting comorbid cases into either the ADHD or ASD sub-phenotype. These analyses were conducted both including and excluding individuals with intellectual disability. Please see **Supplemental Table S7** and **Supplemental Figure S5** for an overview of comparisons.

## Data Availability

Aggregated data (i.e. summary statistics) will be made available on the iPSYCH download page (https://ipsych.dk/en/research/downloads/).

## Acknowledgements

The iPSYCH team was supported by grants from the Lundbeck Foundation (R102-A9118, R155-2014-1724, and R248-2017-2003), the EU H2020 Program (Grant No. 667302, “CoCA”), NIMH (1U01MH109514-01 to ADB) and the Universities and University Hospitals of Aarhus and Copenhagen. The Danish National Biobank resource was supported by the Novo Nordisk Foundation. High-performance computer capacity for handling and statistical analysis of iPSYCH data on the GenomeDK HPC facility was provided by the Center for Genomics and Personalized Medicine and the Centre for Integrative Sequencing, iSEQ, Aarhus University, Denmark (grant to ADB). Dr. Valodakis was funded by NIMH grant K08MH122911 and the 2020 NARSAD Young Investigator Grant #29350 from the Brain & Behavior Research Foundation. Support for this research was further received by Dr. Cormand from the Spanish ‘Ministerio de Ciencia, Innovación y Universidades’ (RTI2018-100968-B-100), ‘Ministerio de Economía y Competitividad’, ‘AGAUR/Generalitat de Catalunya’ (2017-SGR-738), the European Union H2020 Program [H2020/2014-2020] under grant agreements n°667302 (CoCA), 643051 (MiND) and 728018 (Eat2beNICE), and the ECNP network “ADHD across the lifespan”. Dr. Faraone is supported by the European Union’s Horizon 2020 research and innovation programme under grant agreements No 667302 and 965381; NIMH grants U01MH109536-01, U01AR076092-01A1, R0MH116037 and 5R01AG06495502; Oregon Health and Science University, Otsuka Pharmaceuticals and Supernus Pharmaceutical Company.

The authors would like to deeply thank all participants in the cohorts included in this analysis.

## Author contributions

MM, JG and ADB designed the study. MM, JG, TDA, JM, GV, SM, DD, JB, RW, CEC, AR, NS, MEH, BZ and GH conducted data analysis. PBM, EBR, PR, BMN, MJD and ADB supervised data analysis. JB-G, MBH, EA, MN, TW, OM, DMH, PBM, BMN, MJD and ADB provided data. MM, JG, TDA, JM, SM and ADB wrote the paper. MM, JG, TDA, JM, BC, EBR, SVF, BF, SD and ADB comprised the core revision group. ADB directed the study. All authors discussed the results and approved the final version of the manuscript.

