## Supplementary Text and Figures for "Identification of shared and differentiating genetic risk for autism spectrum disorder, attention deficit hyperactivity disorder and case subgroups"

**Supplemental Material and Methods**

#### Functional characterization and annotation of main findings

We used the FUMA v1.3.6a [1] website (http://fuma.ctglab.nl) for downstream functional characterization and annotation of our findings. For all analyses mentioned in the manuscript default settings were applied. More detailed information on available datasets and analytical approaches are available on the website for FUMA (https://fuma.ctglab.nl/tutorial). Please also see Supplemental Table S8 for more details on respective default settings.

##### *eQTL mapping*

For eQTL mapping the following datasets available in FUMA were included

(https://fuma.ctglab.nl/tutorial#eQTLs):

eQTLcatalogue/BrainSeq_ge_brain.txt.gz,
PsychENCODE/PsychENCODE_eQTLs.txt.gz,
scRNA_eQTLs/PBMC.txt.gz,
CMC/CMC_SVA_cis.txt.gz, CMC/CMC_SVA_trans.txt.gz, CMC/CMC_NoSVA_cis.txt.gz,
CMC/CMC_NoSVA_trans.txt.gz,
BRAINEAC/CRBL.txt.gz, BRAINEAC/FCTX.txt.gz, BRAINEAC/HIPP.txt.gz, BRAINEAC/MEDU.txt.gz,
BRAINEAC/OCTX.txt.gz, BRAINEAC/PUTM.txt.gz,
BRAINEAC/SNIG.txt.gz, BRAINEAC/TCTX.txt.gz, BRAINEAC/THAL.txt.gz,
BRAINEAC/WHMT.txt.gz, BRAINEAC/aveALL.txt.gz,
GTEx/v8/Cells_EBV-transformed_lymphocytes.txt.gz, GTEx/v8/Whole_Blood.txt.gz,
GTEx/v8/Brain_Amygdala.txt.gz, GTEx/v8/Brain_Anterior_cingulate_cortex_BA24.txt.gz,
GTEx/v8/Brain_Caudate_basal_ganglia.txt.gz, GTEx/v8/Brain_Cerebellar_Hemisphere.txt.gz,
GTEx/v8/Brain_Cerebellum.txt.gz, GTEx/v8/Brain_Cortex.txt.gz, GTEx/v8/Brain_Frontal_Cortex_BA9.txt.gz,
GTEx/v8/Brain_Hippocampus.txt.gz,
GTEx/v8/Brain_Hypothalamus.txt.gz, GTEx/v8/Brain_Nucleus_accumbens_basal_ganglia.txt.gz,
GTEx/v8/Brain_Putamen_basal_ganglia.txt.gz, GTEx/v8/Brain_Spinal_cord_cervical_c-1.txt.gz,
GTEx/v8/Brain_Substantia_nigra.txt.gz

No filtering (e.g. based on CADD scores or other available information) was applied.

#### *Chromatin Interaction mapping*

For chromatin interaction mapping the following datasets available in FUMA were used (https://fuma.ctglab.nl/tutorial#chromatin-interactions):

EP/PsychENCODE/EP_links_oneway.txt.gz, HiC/PsychENCODE/Promoter_anchored_loops.txt.gz,
HiC/Giusti-Rodriguez_et_al_2019/Adult_Cortex.txt.gz,
HiC/Giusti-Rodriguez_et_al_2019/Fetal_Cortex.txt.gz,
HiC/GSE87112/Dorsolateral_Prefrontal_Cortex.txt.gz,
HiC/GSE87112/Hippocampus.txt.gz,
HiC/GSE87112/Neural_Progenitor_Cell.txt.g

Again, no posterior filtering was applied.

#### *Single Cell Analyses*

General details for single cell analyses within the FUMA framework can be found on the developer's website (https://fuma.ctglab.nl/tutorial#celltype). We used three of the available datasets within FUMA (https://fuma.ctglab.nl/tutorial#datasets):

PsychENCODE data for human developmental and adult brain samples [2].
GSE76381 data for human brain samples (ventral midbrain from 6-11 weeks embryos) [3].

For naming conventions on different cell types used in the three datasets please see the original publications [2,3]. In brief for the PsychENCODE data: *Ex1 to Ex9* and *In1 to In8* - excitatory and inhibitory neurons; *OPC* - oligodendrocyte progenitor cells, *IPC - intermediate progenitor cells; NEP* - neuroepithelial cells; *trans* - transient cell type. For GSE76381: *DA0-2* - dopaminergic neurons; *Endo* - endothelial cells; *Gaba* - GABAergic neurons; *Mgl* - microglia; *NProg* - neuronal progenitor; NbGaba - neuroblast gabaergic; *NbM* - medial neuroblast; *NbML1+5* - mediolateral neuroblasts; *OMTN* - oculomotor and trochlear nucleus; *OPC* - oligodendrocyte precursor cells. *Peric* - pericytes; *Prog* - progenitor medial floorplate (FPM), lateral floorplate (FPL), midline (M), basal plate (BP); *RN* - red nucleus; *Rgl1-3* - radial glia-like cells; *Sert* – serotonergic.

### Supplemental Figure S1: Regional association plot for combined meta-analysis.

a) rs7538463

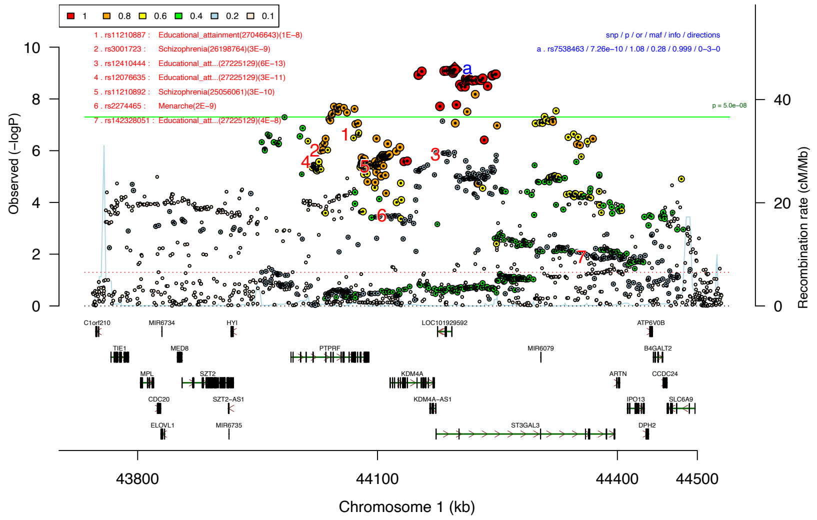

b) rs4916723

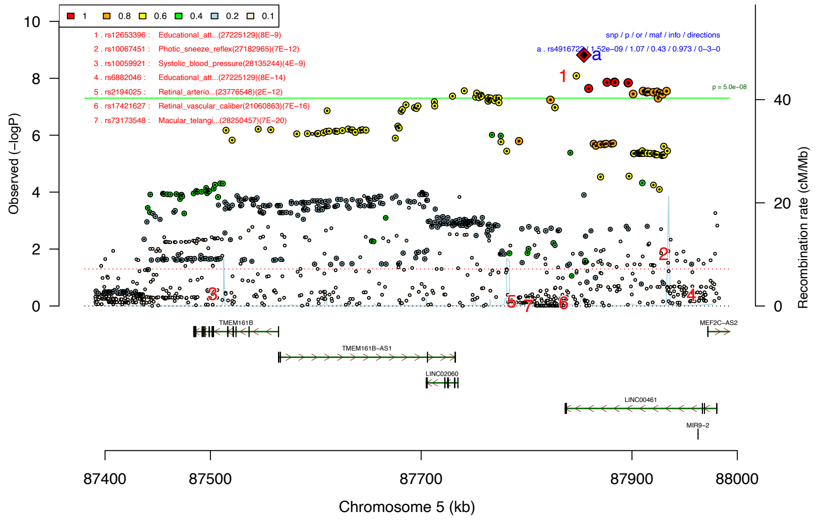

c) rs2391769

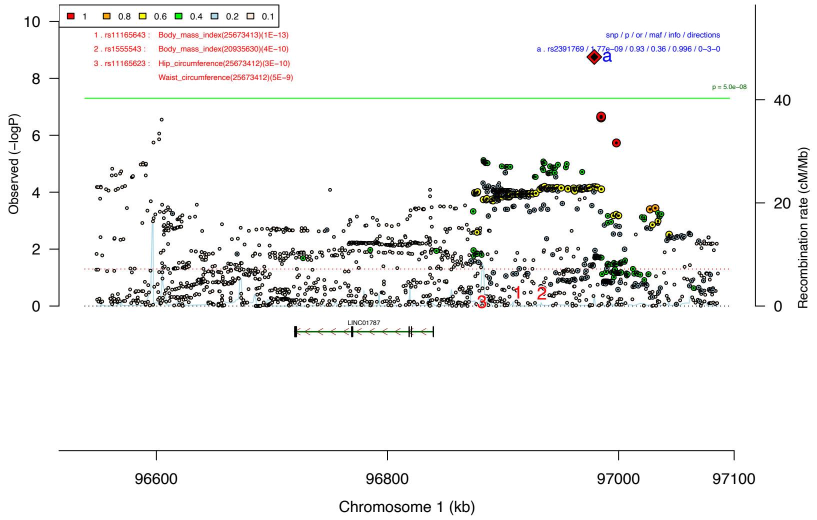

d) rs9530773

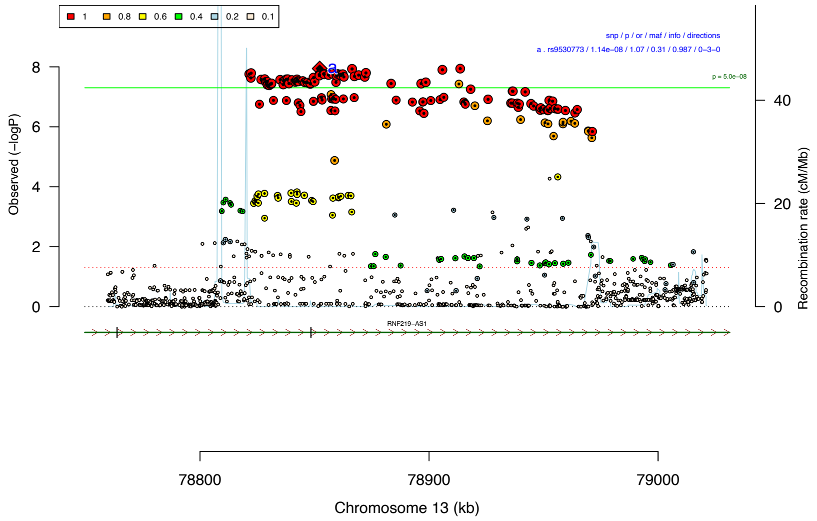

e) rs138696645

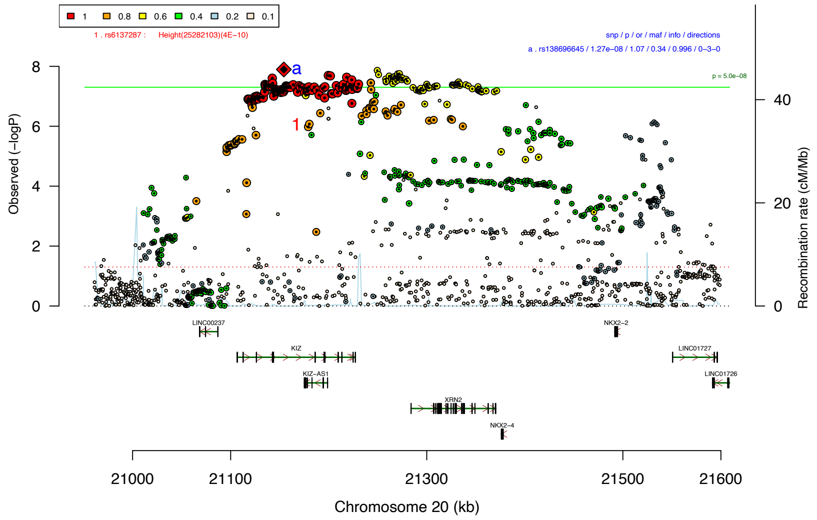

f) rs227293

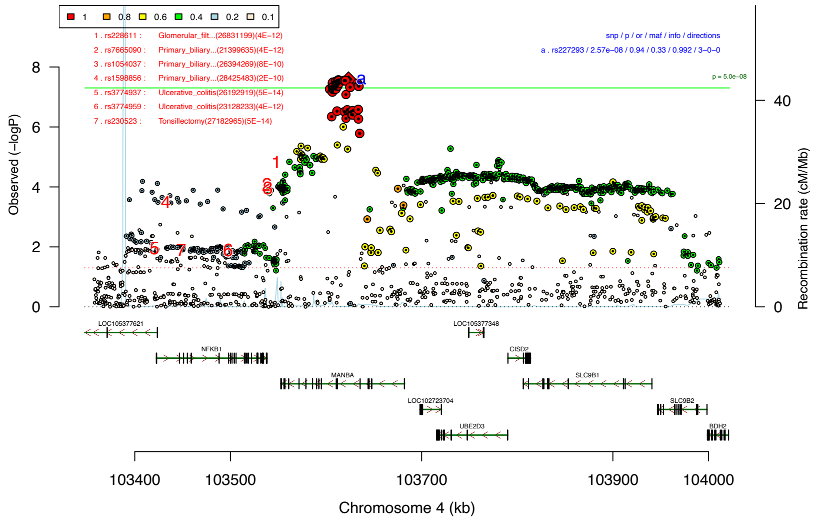

g) rs325506

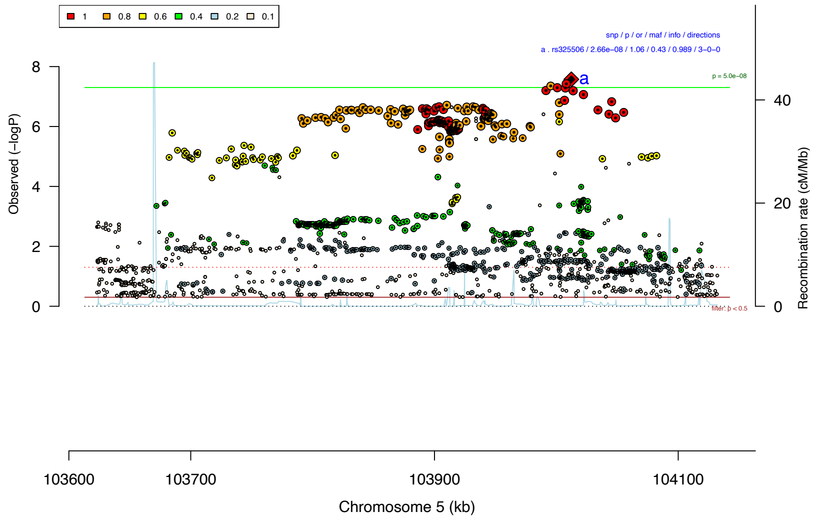

Regional association plots showing association significances for the top seven linkage disequilibrium (LD)-independent index SNPs and all markers within a region of strong LD. SNPs are color coded according to strength of LD with respect to lead SNP (black diamond with red corners) in each region (defined by $r^2$ statistic). Estimated recombination rates from HapMap phase3 CEU reference panel are depicted as blue lines along the physical position of each region. Genes are drawn in the bottom quarter of the plot (unless in a region devoid of genes) with vertical bars denoting positions of exons. LD-independent genome-wide significant hits are labeled with lower case letters and a list of main characteristics is provided (**snp** – marker name, **p** – P-value of association, **or** – Odds ratio for association, **maf** – Minor allele frequency, **info** – INFO score obtained through PLINK for associated marker, directions – brief table of direction of effects). We used data from the GWAS catalog (as of Oct 2017) to annotate region with known GWAS hits (if there are any), please refer to Supplemental Table S1 and **Supplemental Figure S2** for a more detailed overview. In the annotations, numbers are used to highlight previously associated markers within the plot and a corresponding table is provided. In one of the regional association plots **(g)** only SNPs below a P-value ($p < 0.5$) are shown, in all other instances all SNPs in the region are plotted.

#### Supplemental Figure S2: PheWAS plots for associated SNPs from combined GWAS

a) rs7538463

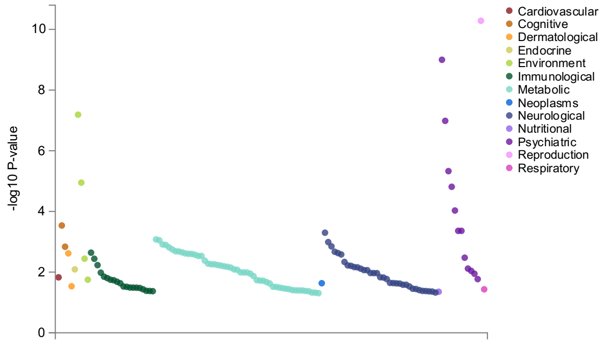

b) rs4916723

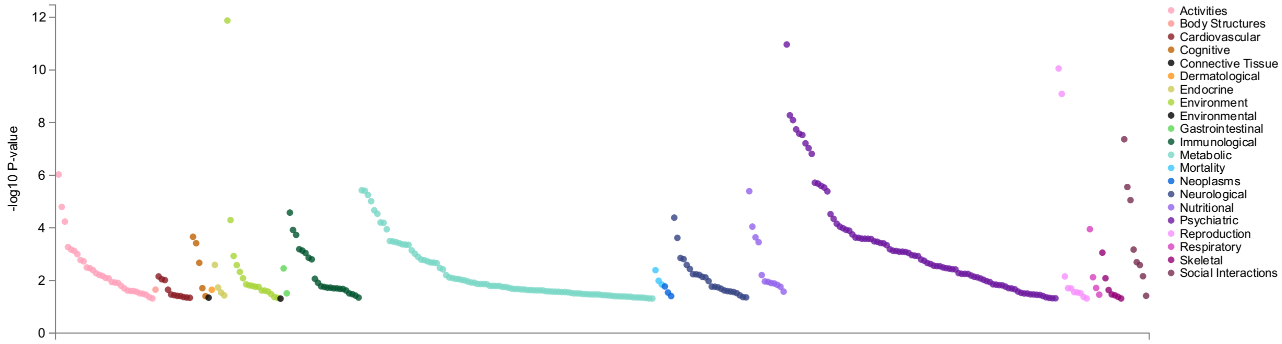

c) rs2391769

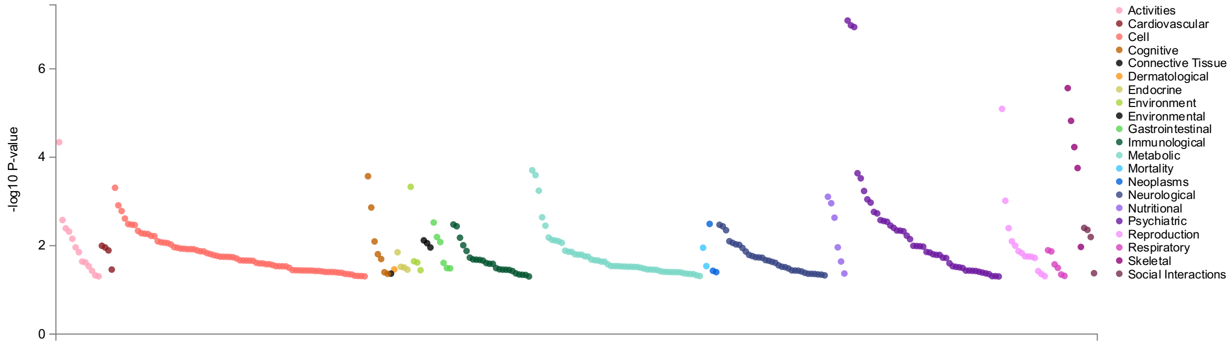

d) rs9530773

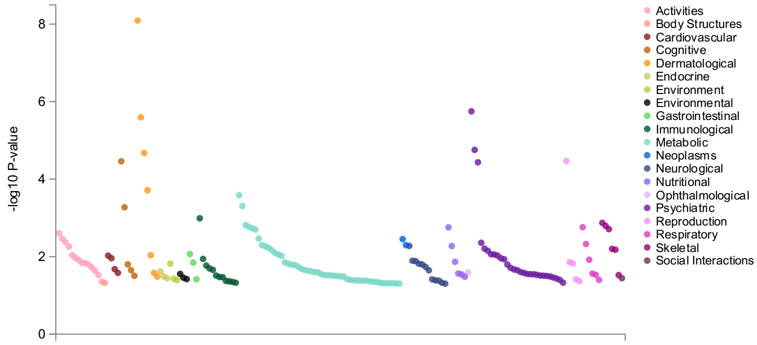

e) rs138696645

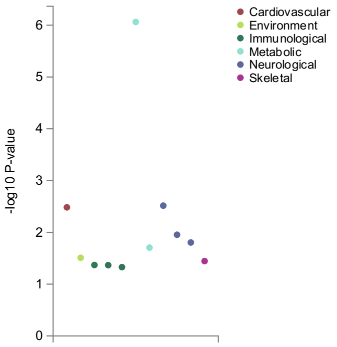

f) rs227293

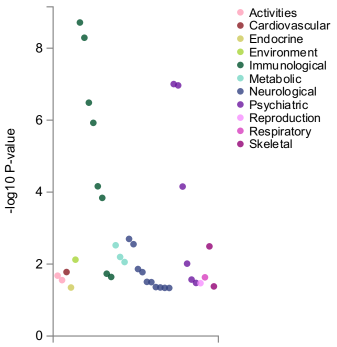

g) rs325506

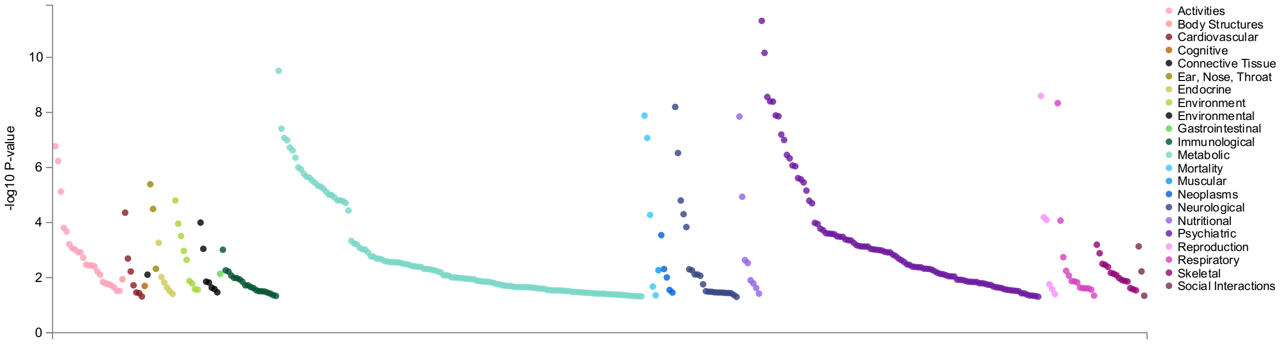

PheWAS analyses with gwasATLAS [4]. Default p-value cutoff at 0.05, traits ordered by domain and p-value. Overall number of GWASs considered for these analyses: 4,756. This also includes GWASs in which the searched SNP was not tested (Bonferroni corrected P-value: $p = 1.05^{x}10^{-5}$).

#### Supplemental Figure S3: Regional Miami plots for combined and ADHD vs ASD GWASs.

a) KRT8P46 (chromosome 4; transcript: KRT8P46-201)

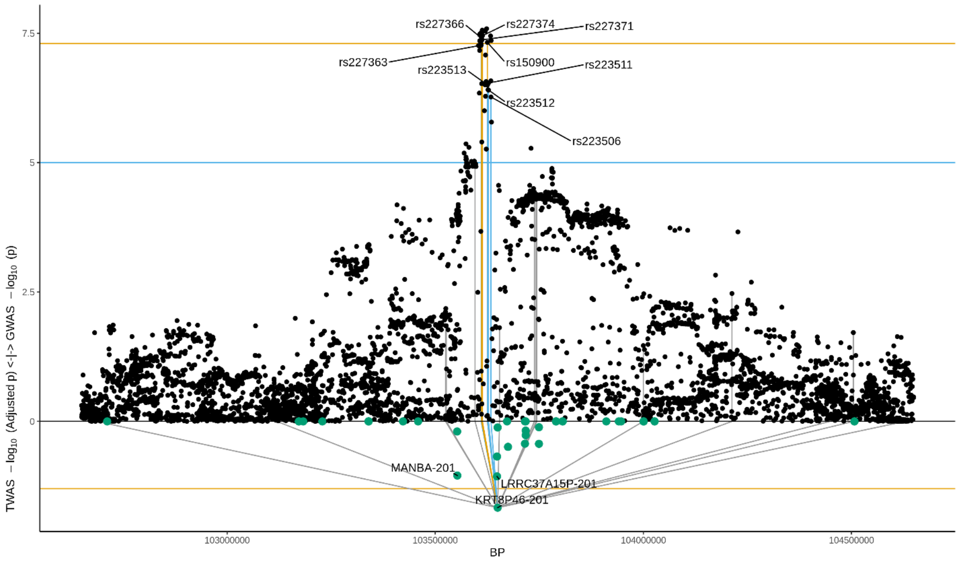

b) HIST1H2BD (chromosome 6; transcript: HIST1H2BD-201)

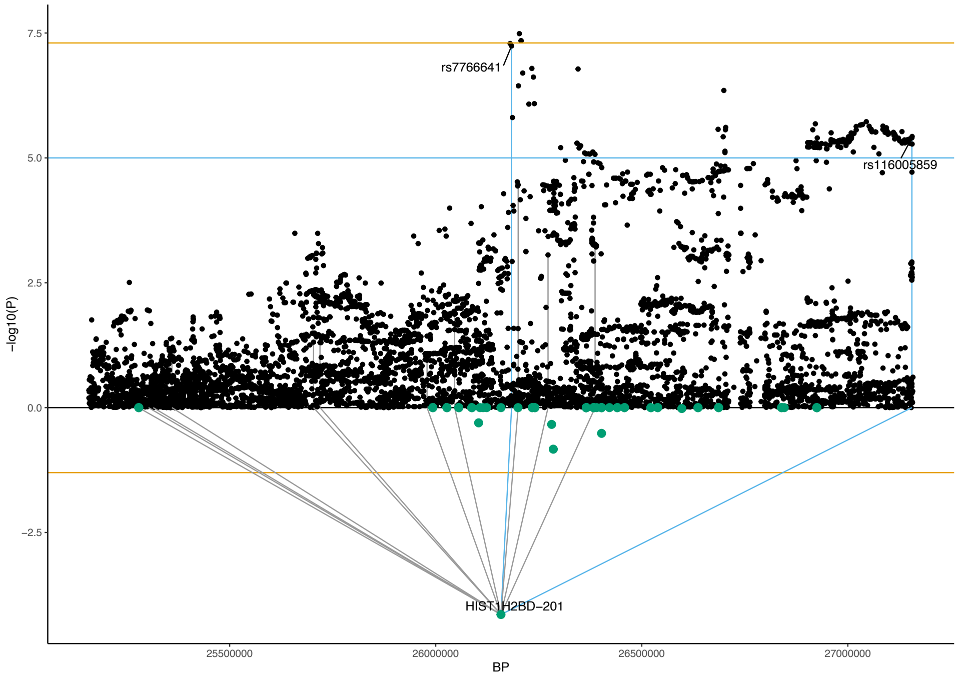

c) CAMKV (chromosome 3; transcript: CAMKV-210)

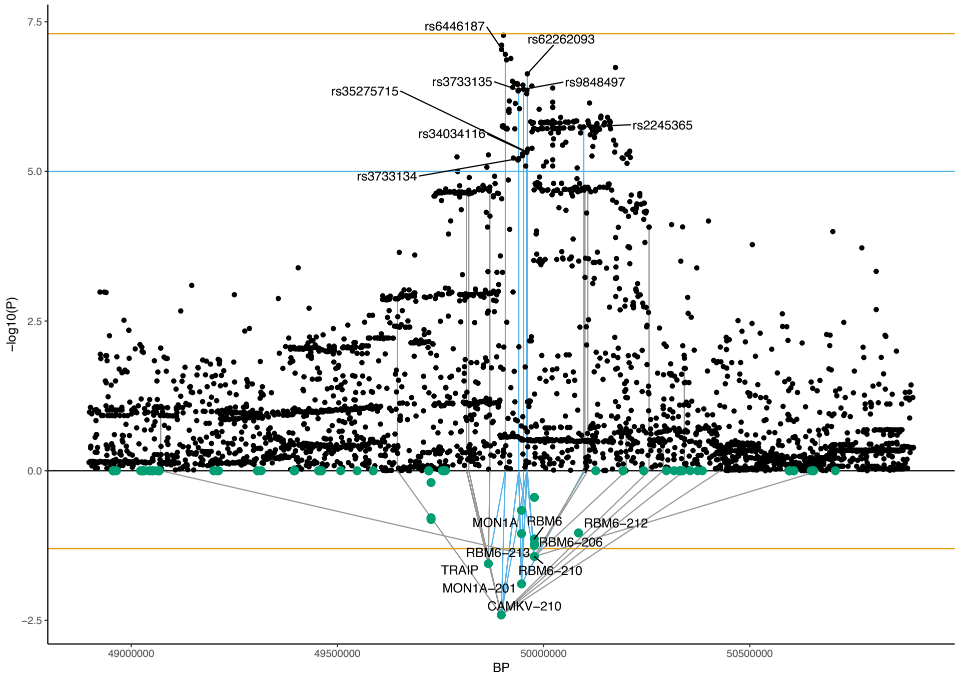

d) RFT1 (chromosome 3; transcript: RFT1-204)

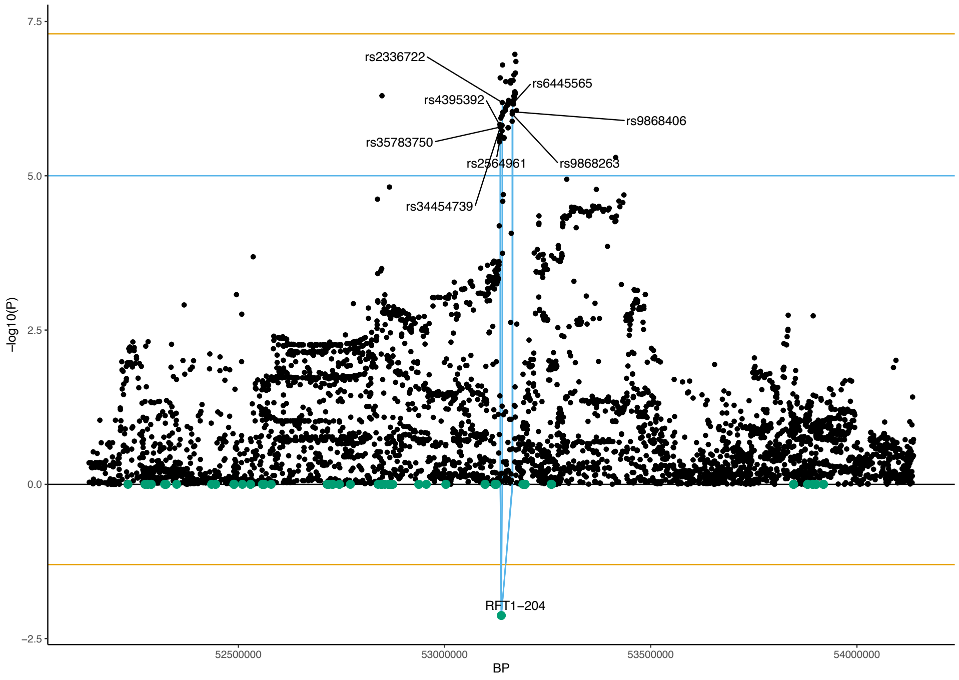

e) FAM167A (chromosome 8; gene)

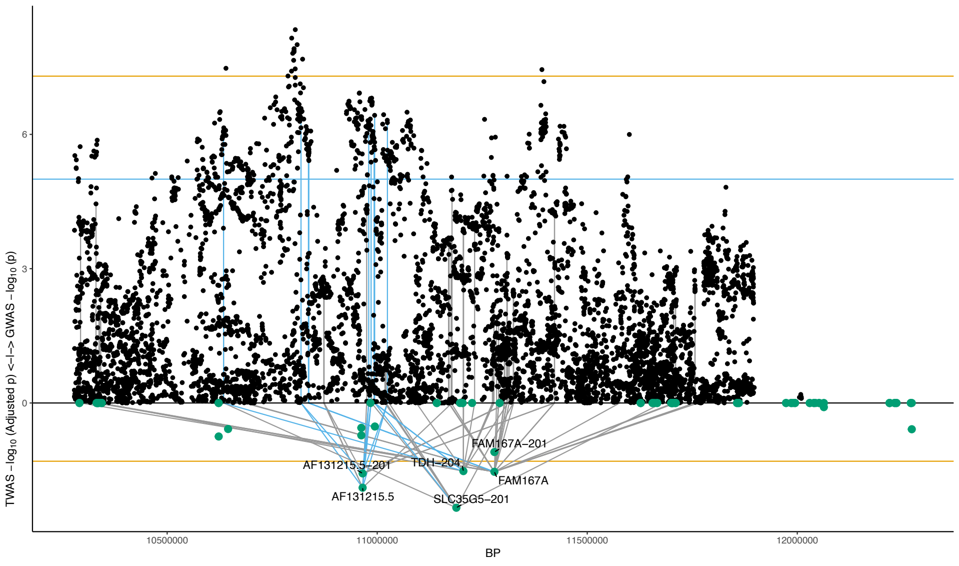

Regional Miami plots for **(a)** combined and **(a-d)** ADHD vs ASD GWASs corresponding to the genomic region of the respective transcript (1Mbp window from start site). Please also refer to Supplemental Table S2 for details. **Top panel** shows the GWAS results (***black dots***); ***blue line*** corresponds to $p = 1^x10^{-5}$, ***orange line*** to $p = 5^x10^{-8}$ (genome-wide significance). **Bottom panel** shows the TWAS results (***green dots***; only the transcripts with Bonferroni-adjusted $p < 0.1$ are labelled for clarity) for different transcripts (genes are represented by both gene expression and isoform expression); ***orange line*** corresponds to Bonferroni-adjusted $p = 0.05$. Each transcript that is Bonferroni-significant in the region is connected with lines to the SNPs that contribute to its transcriptomic imputation model; lines are ***grey*** when the SNPs have a $p > 1^x10^{-5}$, ***blue*** when $p < 1^x10^{-5}$ but $> 5^x10^{-8}$ and ***orange*** when $p < 5^x10^{-8}$. The SNPs that are above the blue line and contribute to the transcriptomic imputation models of significant transcripts are labelled.

#### Supplemental Figure S4: Manhattan Plot for gene-based analyses in main GWAS comparisons.

a)

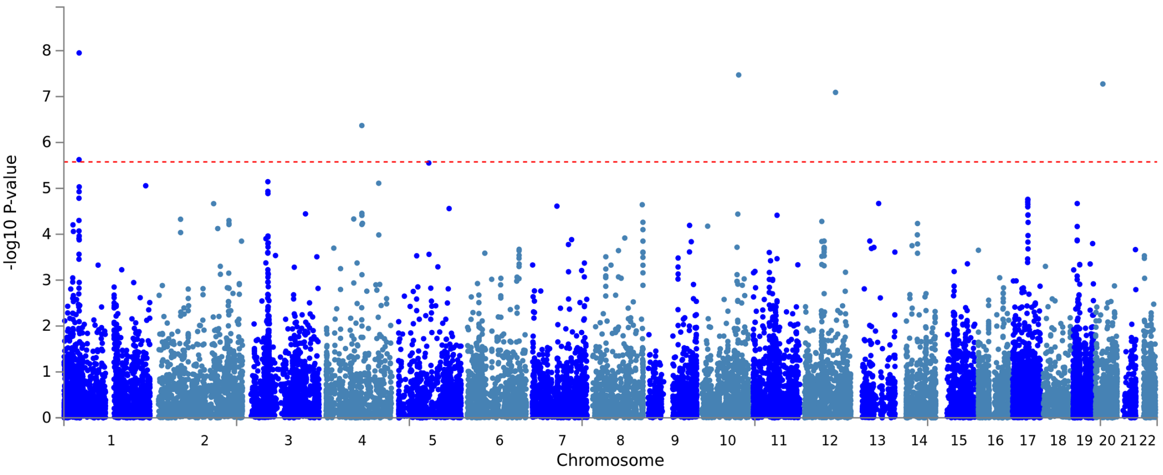

b)

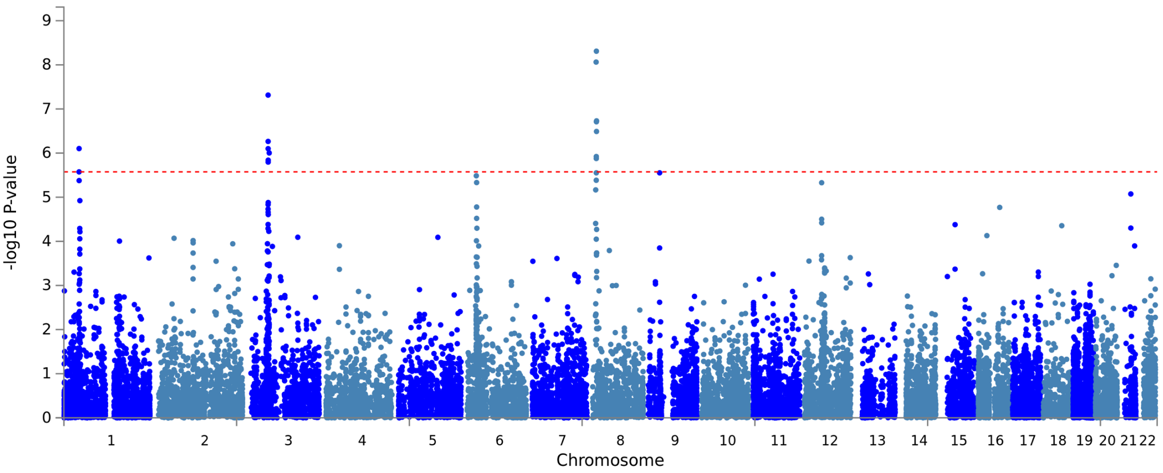

Results from analyses using MAGMA v 1.08 [5] with default settings (and without using a padding sequence) as implemented in FUMA [1]. The x-axis in both sub-plots shows the position in the genome (chromosomes 1–22) and the y-axis the statistical significance as –log10 (P). Red line indicates genome-wide significance. **(a)** Results of genome-wide analyses for combined GWAS (34,462 cases and 41,201 controls). Each dot represents one of the 18,837 genes tested in the analysis. Two of the genes (*SORCS3* and *DUSP6*) are located in regions that were not identified in the GWAS, suggesting these as additional shared loci. **(b)** Results of genome-wide analyses for ADHD vs ASD GWAS (11,964 ADHD only cases and 9,315 ASD only cases). Each dot represents one of the 18802 genes tested in the analysis. There were 14 genome-wide significant ($p < 2.66^x10^{-6}$) gene-based associations detected. Nine of these genes are novel associations not previously identified in gene-wise analyses of the disorders separately in the largest GWASs published to date [6,7] (Supplemental Table S3). However, three of the 9 genes are located in the chromosome 8 region that also harbors *SOX7*, *XRK6*, and *BLK*, genes that have been found associated with ASD before [6]. The remaining six genes are located on chromosome 3 in a locus that was not genome-wide significant in the GWAS.

### Supplemental Figure S5: Regional association plots for ADHD vs ASD GWAS.

a) rs13023832

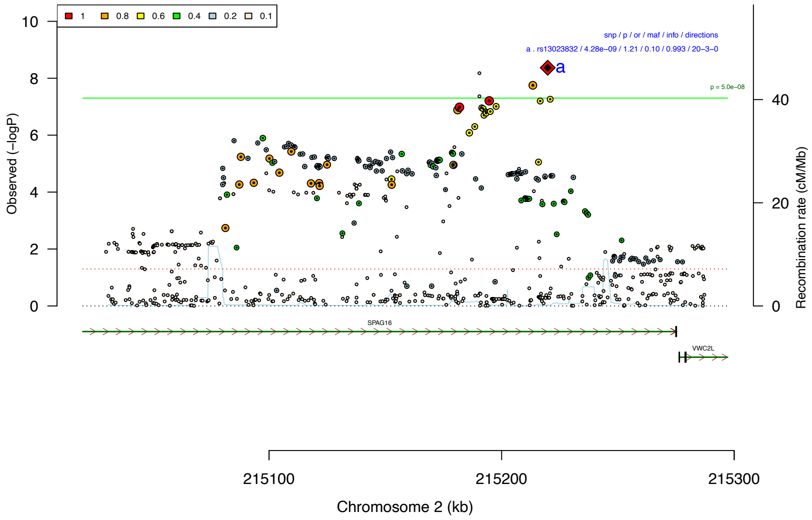

b) rs7821914

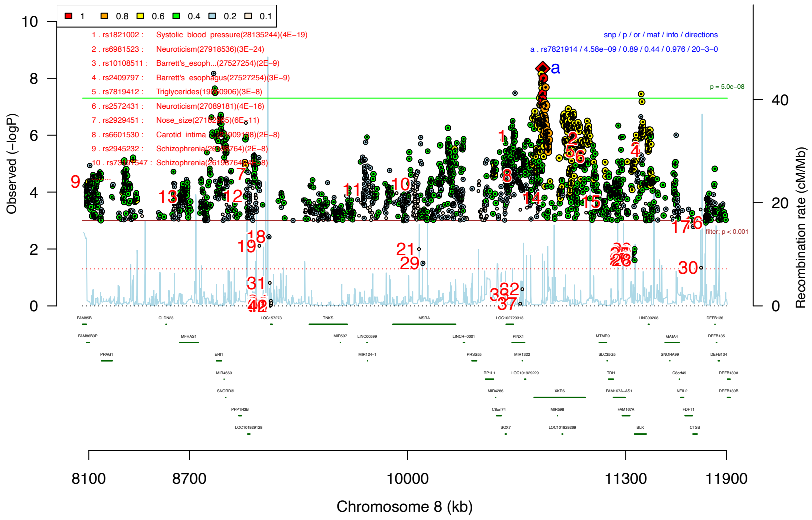

c) rs147420422

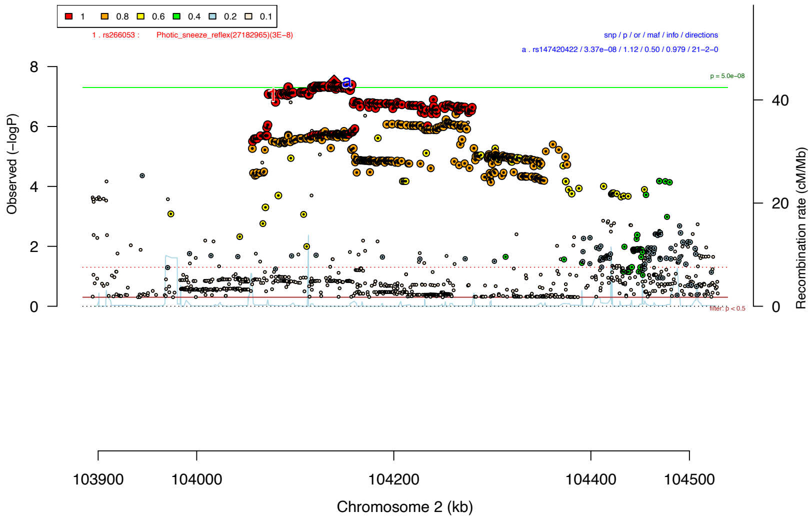

d) rs3791033

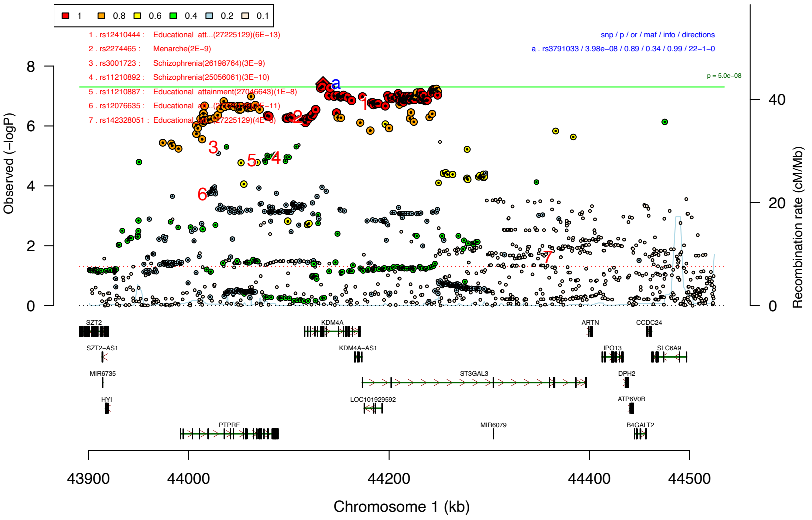

e) rs9379833

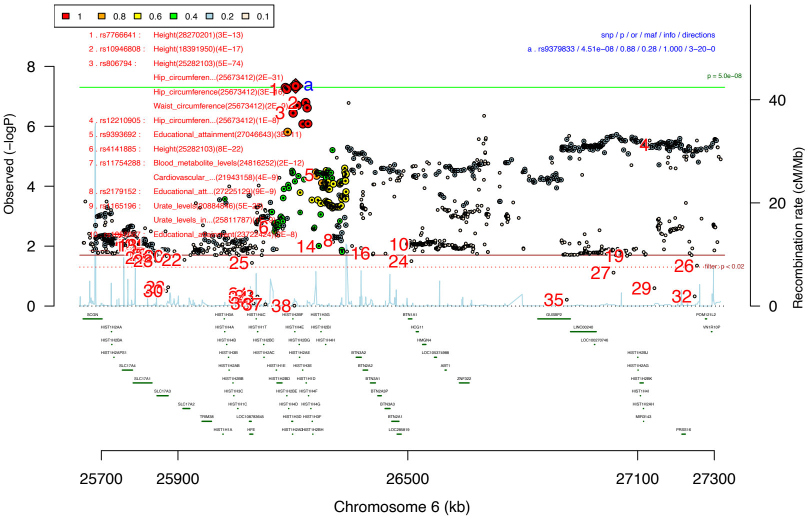

Regional association plots showing association significances for the top five linkage disequilibrium (LD)-independent index SNPs and all markers within a region of strong LD. SNPs are color coded according to strength of LD with respect to lead SNP (black diamond with red corners) in each region (defined by $r^2$ statistic). Estimated recombination rates from HapMap phase3 CEU reference panel are depicted as blue lines along the physical position of each region. Genes are drawn in the bottom quarter of the plot (unless in a region devoid of genes) with vertical bars denoting positions of exons. LD-independent genome-wide significant hits are labeled with lower case letters and a list of main characteristics is provided (snp – marker name, p – P-value of association, or – Odds ratio for association, maf – Minor allele frequency, info – INFO score obtained through PLINK for associated marker, directions – brief table of direction of effects). We used data from the GWAS catalog (as of Oct 2017) to annotate region with known GWAS hits (if there are any), please refer to Supplemental Table S5 and **Supplemental Figure S6** for a more detailed overview. In the annotations, numbers are used to highlight previously associated markers within the plot and a corresponding table is provided. In one of the regional association plots **(b)** only SNPs below a P-value ($p < 0.001$) are shown, or if they have beenb previously identified to be associated with a trait listed in the GWAS catalog. In regional association plot **(c)** only SNPs below a P-value ($p < 0.5$) are shown, in regional association plot **(e)** only SNPs below a P-value ($p < 0.02$) are shown and regional association plots **(a)** and **(d)** all SNPs in the region are plotted.

#### Supplemental Figure S6: PheWAS plots for associated SNPs from ADHD vs ASD GWAS

a) rs13023832

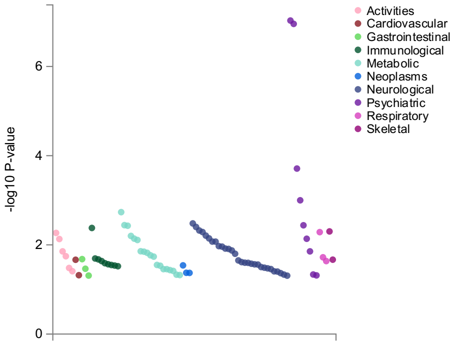

b) rs7821914

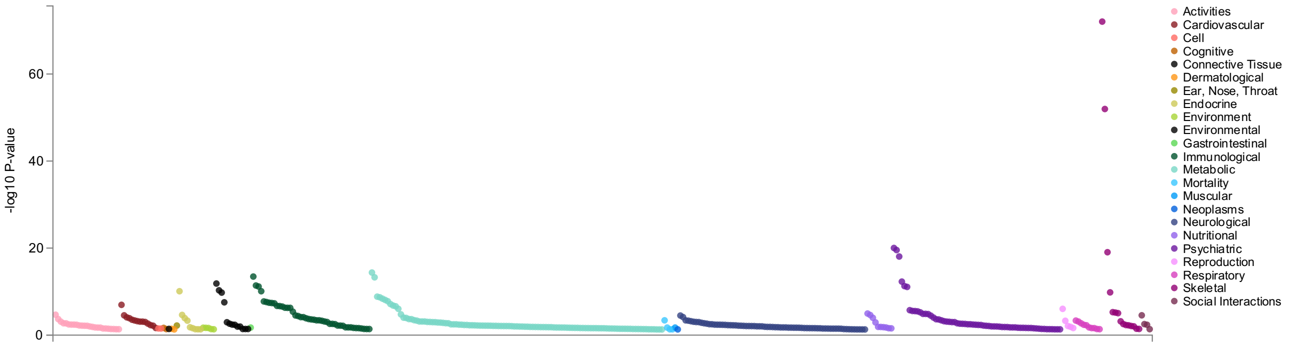

c) rs147420422

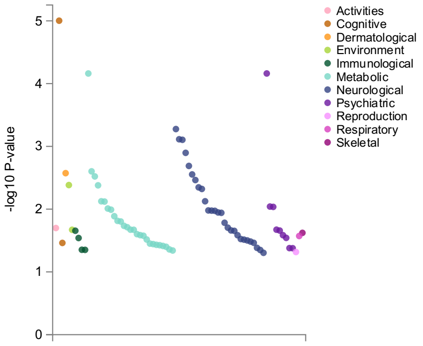

d) rs3791033

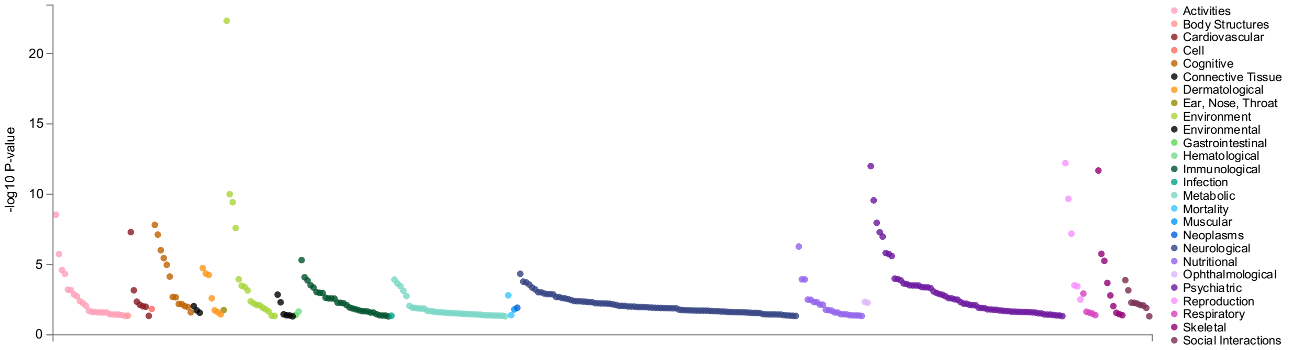

e) rs9379833

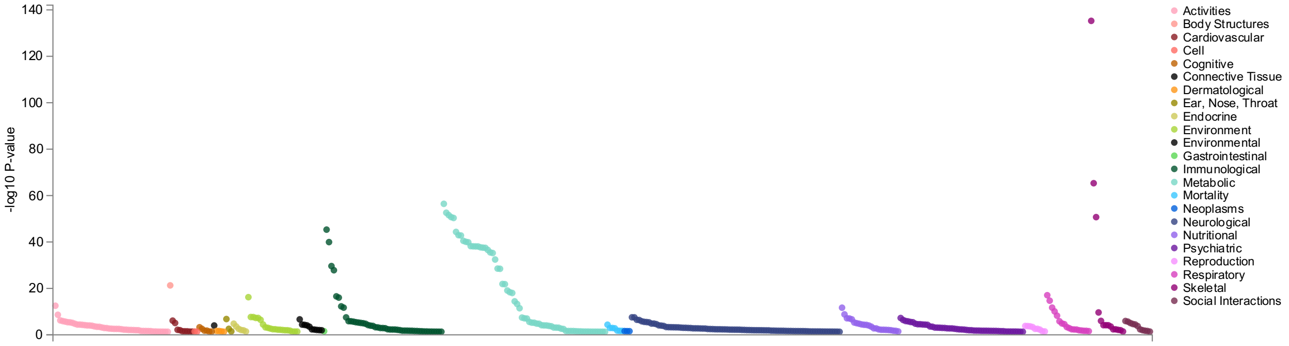

PheWAS analyses with gwasATLAS [4]. Default p-value cutoff at 0.05, traits ordered by domain and p-value. Overall number of GWASs considered for these analyses: 4,756. This also includes GWASs in which the searched SNP was not tested (Bonferroni corrected P-value: $p = 1.05^{x}10^{-5}$).

#### Supplemental Figure S7: Genetic correlations between ADHD and ASD with other traits and disorders.

a)

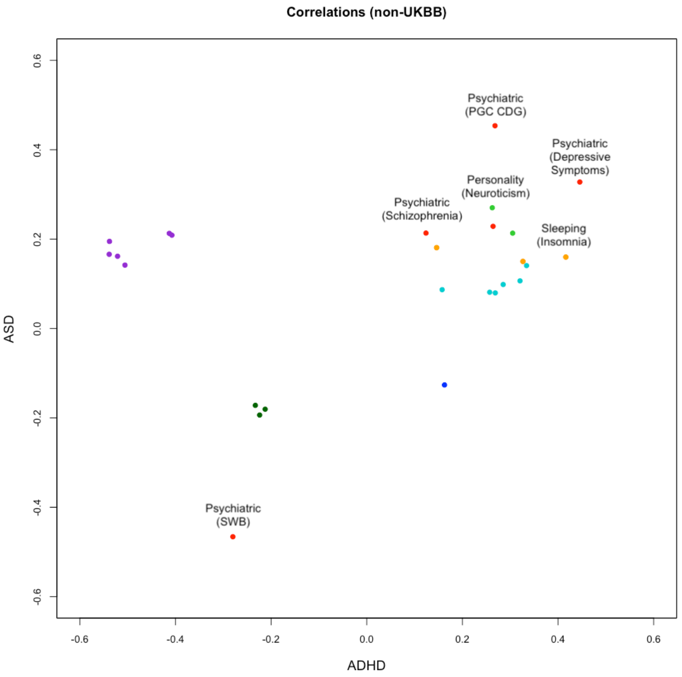

b)

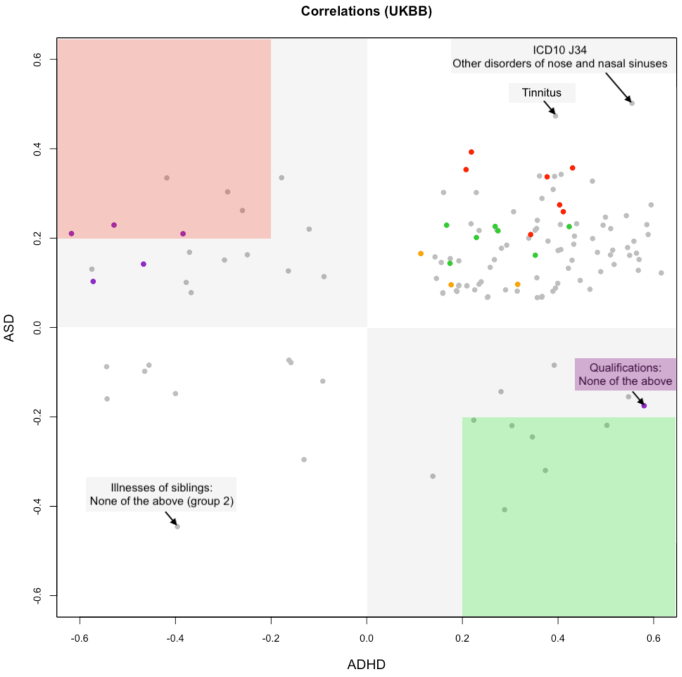

Genetic correlations for ADHD (PMID 30478444) and ASD (PMID 30804558) with other traits as calculated by LDhub (PMID 27663502). Please refer to Supplemental Table S4 for an overview of all genetic correlations. In **(a)** "Non-UKBB" (*n*= 28) and **(b)** "UKBB" (*n*= 136) only correlations with a *Z score* > 2 in both ADHD and ASD are shown ("*pass*" in Supplemental Table S4 – OVERVIEW for *z2_flag_asd* and *z2_flag_adhd*; *n*= 164 in total). **(a)** Non-UKBB: Color is coded as follows for different categories (please note that results for cross correlations for ADHD or ASD GWASs are not shown): personality (Neuroticism with two GWASs) – green, psychiatric (*Subjective well-being (SWB); PGC cross-disorder analysis (PGC CDG); Depressive Symptoms, Schizophrenia, Major Depressive Disorder*) – red, education/ cognitive (*Years of schooling (proxy cognitive performance), Years of schooling 2016; Years of schooling 2013; College completion; Childhood IQ; Intelligence*) – purple, autoimmune (*Rheumatoid Arthritis*) – blue, sleeping (*2 Insomnia GWASs; Excessive daytime sleepiness*) – orange, anthropometric (*3 Obesity GWASs (class 1, 2, 3); Waist circumference; Body Mass Index; Hip circumference*) – turquoise, metabolites (*Concentration of large HDL particles; Phospholipids in large HDL; Total cholesterol in HDL*) – dark green. **(b)** UKBB: Areas are shaded in grey if they contain rGs for ADHD and ASD that are positive for one and negative for the other. The areas are colored in light reen if they have a rG > 0.2 for ADHD and one for ASD < -0.2 and in light red if they have a rG < -0.2 for ADHD and one for ASD > 0.2. There are 6 UKBB traits in either of the two colored areas: *light green* - *Job involves mainly walking or standing* (ADHD: 0.50, ASD: -0.22), *Transport type for commuting to job workplace: Car/motor vehicle* (0.37, -0.32), *Weight change compared with 1 year ago* (0.35, -0.24), *Duration of vigorous activity* (0.30, -0.22), *Number of children fathered* (0.29, -0.41), *Prospective memory result* (0.22, -0.21); *light red* - *Qualifications: A levels/AS levels or equivalent* (ADHD: -0.62, ASD 0.21), *Qualifications: College or University degree* (-0.53, 0.23), *Transport type for commuting to job workplace: Public transport* (-0.42, 0.34), *Fluid intelligence score* (-0.38, 0.21), *Types of transport used (excluding work): Public transport* (-0.29, 0.30), *Transport type for commuting to job workplace: Walk* (-0.26, 0.26). personality (*Frequency of tenseness / restlessness in last 2 weeks, Fed-up feelings, Guilty feelings, Tense / highly strung, Irritability, Sensitivity / hurt feelings, Neuroticism score*) – green, psychiatric (*Number of depression episodes, Loneliness_ isolation, Illnesses of siblings: Severe depression, Miserableness, Frequency of depressed mood in last 2 weeks, Ever unenthusiastic/disinterested for a whole week, Ever depressed for a whole week*) – red, education/ cognitive (*Qualifications: A levels/AS levels or equivalent, Qualifications: College or University degree, Qualifications: None of the above, Qualifications: O levels/GCSEs or equivalent, Qualifications: Other professional qualifications e.g.: nursing_ teaching, Fluid intelligence score*) – purple, sleeping (*Sleeplessness / insomnia, Nap during day, Daytime dozing / sleeping (narcolepsy)*) – orange.

**Supplemental Figure S8 MAGMA tissue expression analysis for combined GWAS.**

a)

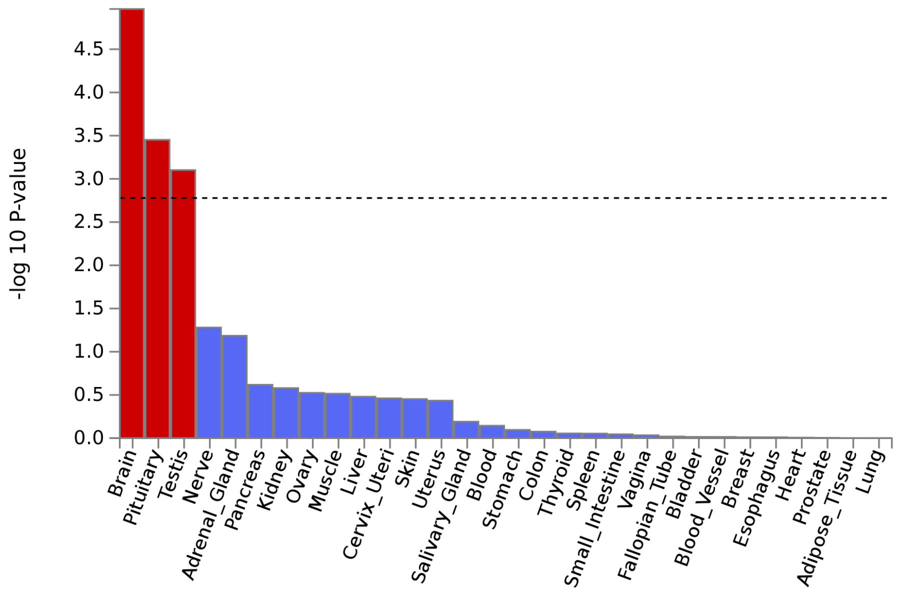

b)

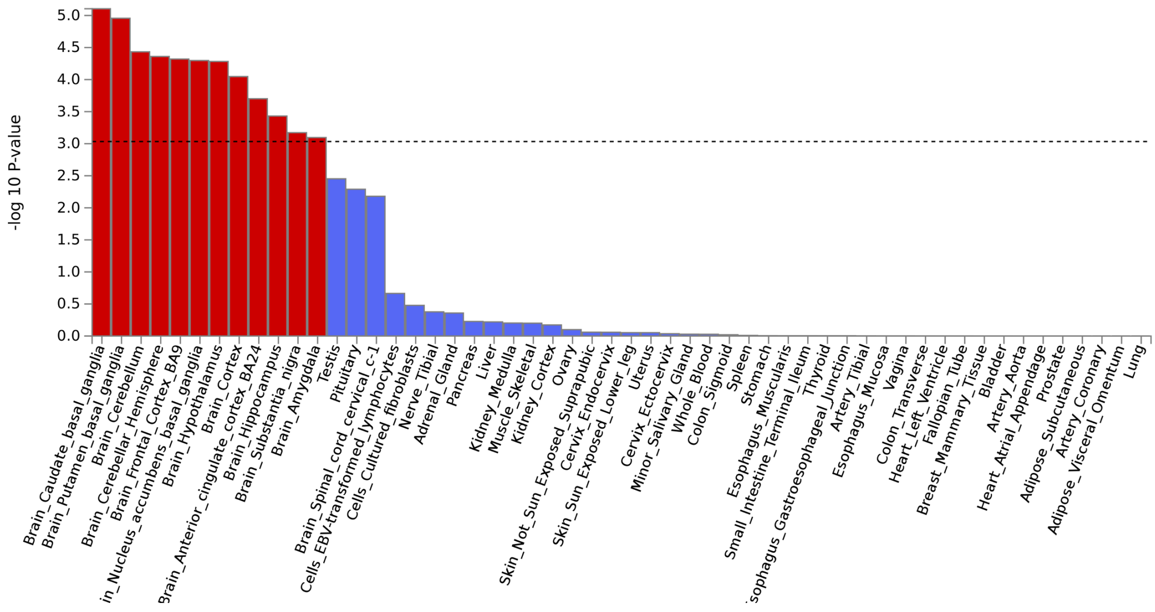

Results of gene-property analysis in MAGMA [5] as implemented in FUMA [1]. Tissue specific data is obtained from the GTEx v8 dataset (https://www.gtexportal.org) [8]. Tissues with red bar surpass experiment-wide significance. **(a)** General tissue type analysis with 30 tissues. **(b)** Individual tissue type analyses with 54 tissue types.

**Supplemental Figure S9 FUMA single-cell analyses for Combined GWAS.**

a)

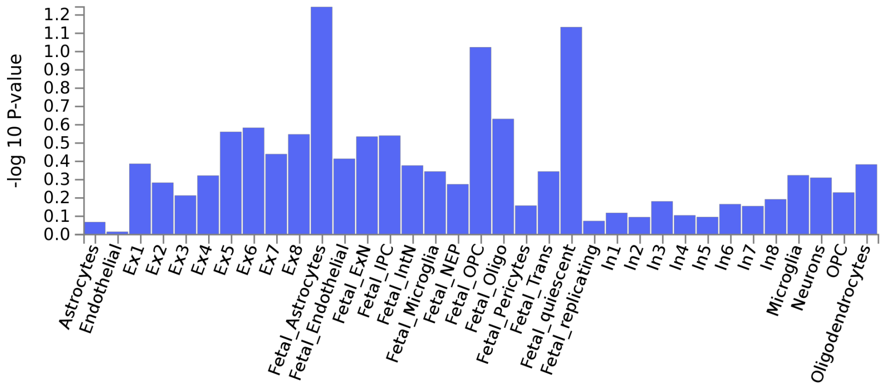

b)

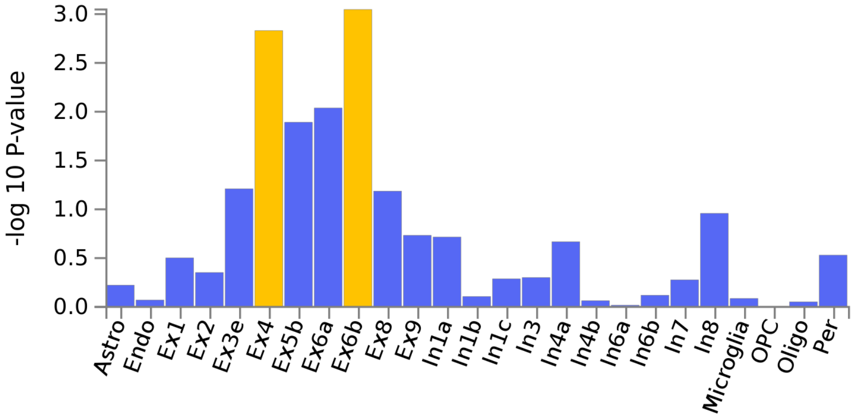

c)

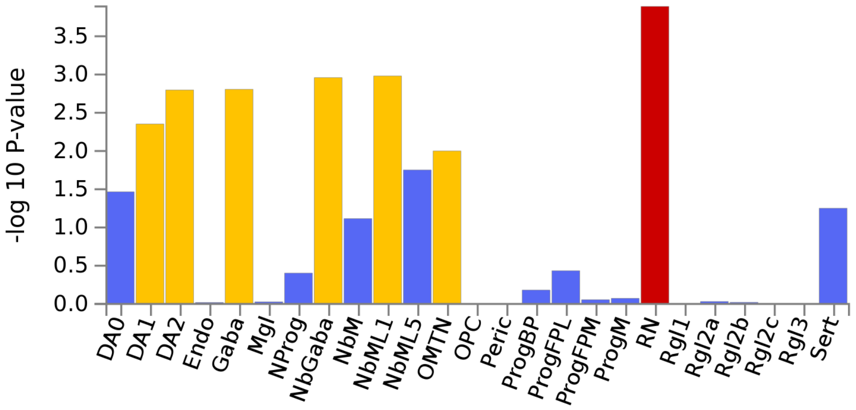

Results are show for the psychENCODE datasets [2] with **(a)** human developmental and **(b)** human adult brain samples as well as the human midbrain cell types (ventral midbrain from 6-11 weeks embryos) from La Manno et al. [3] **(c**, GSE76381). For psychENCODE developmental dataset, 4,249 cells were available, for the psychENCODE adult dataset, 27,380 cells were included in the analysis. Mapping to unique ENSG IDs was available for 15,019 and 16,243 genes, respectively. For the human midbrain samples 1,695 cells were used. Mapping to unique ENSG ID was available for 16,885 genes. Cell types with blue bars are not significant, those in yellow are significant within the tested category, those in red are significant after correction for multiple testing across categories. For naming conventions on different cell types please see the original publication [PMID 30545857 27716510]. In brief for the ***PsychENCODE*** data: *Ex1 to Ex9* and *In1 to In8* - excitatory and inhibitory neurons; *OPC* - oligodendrocyte progenitor cells, *IPC - intermediate progenitor cells; NEP* - neuroepithelial cells; *trans* - transient cell type. For ***GSE76381***: *DA0-2* - dopaminergic neurons; *Endo* - endothelial cells; *Gaba* - GABAergic neurons; *Mgl* - microglia; *NProg* - neuronal progenitor; NbGaba - neuroblast gabaergic; *NbM* - medial neuroblast; *NbML1+5* - mediolateral neuroblasts; *OMTN* - oculomotor and trochlear nucleus; *OPC* - oligodendrocyte precursor cells. *Peric* - pericytes; *Prog* - progenitor medial floorplate (FPM), lateral floorplate (FPL), midline (M), basal plate (BP); *RN* - red nucleus; *Rgl1-3* - radial glia-like cells; *Sert* – serotonergic.

**Supplemental Figure S10 MAGMA tissue expression analysis for ADHD vs ASD GWAS.**

a)

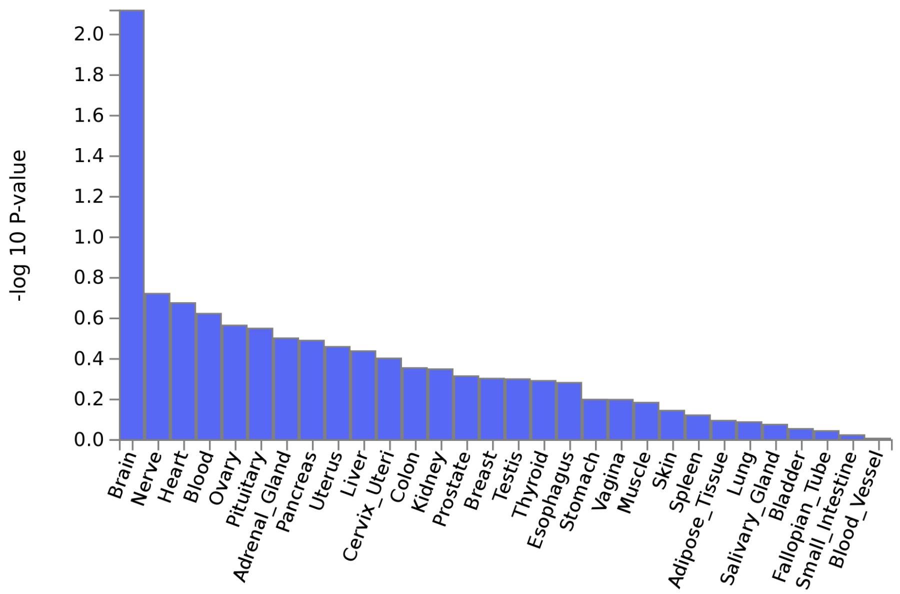

b)

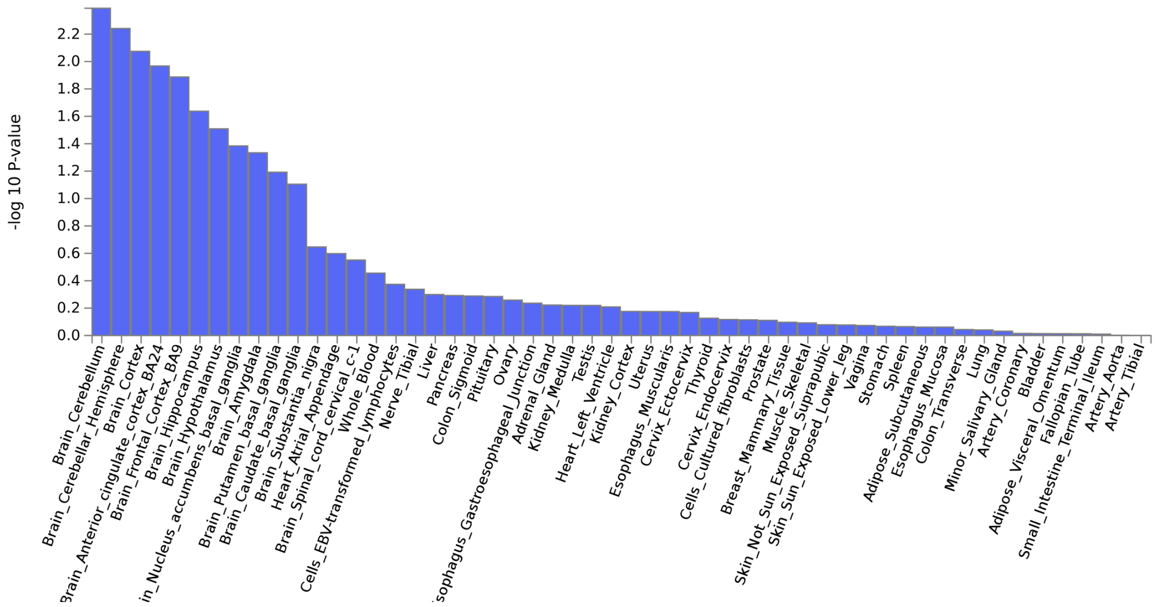

Results of gene-property analysis in MAGMA [5] as implemented in FUMA [1]. Tissue specific data is obtained from the GTEx v8 dataset (https://www.gtexportal.org) [8]. Tissues with red bar surpass experiment-wide significance. **(a)** General tissue type analysis with 30 tissues. **(b)** Individual tissue type analyses with 54 tissue types.

**Supplemental Figure S11 FUMA single-cell analyses for ADHD vs ASD GWAS.**

a)

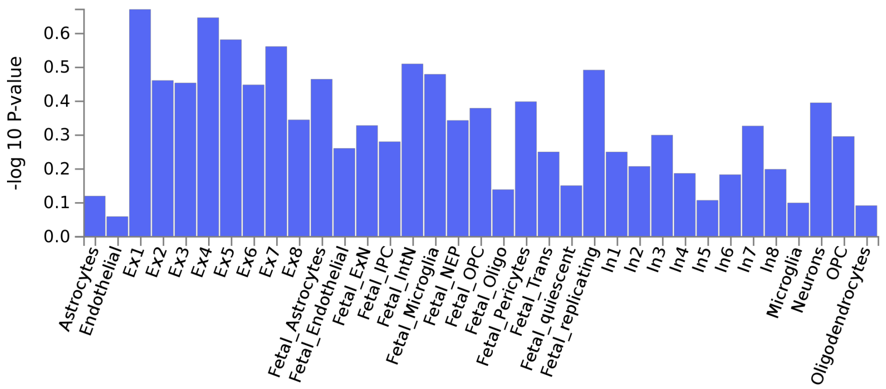

b)

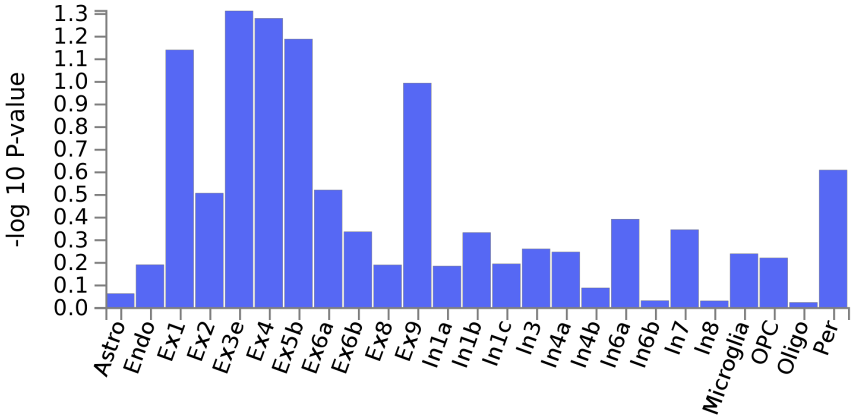

c)

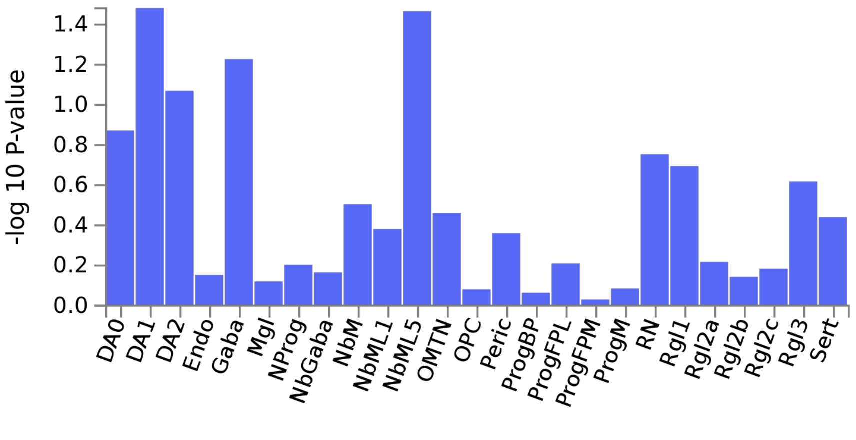

Results are show for the psychENCODE datasets [2] with **(a)** human developmental and **(b)** human adult brain samples as well as the human midbrain cell types (ventral midbrain from 6-11 weeks embryos) from La Manno et al. [3] **(c**, GSE76381). For psychENCODE developmental dataset, 4,249 cells were available, for the psychENCODE adult dataset, 27,380 cells were included in the analysis. Mapping to unique ENSG IDs was available for 15,019 and 16,243 genes, respectively. For the human midbrain samples 1,695 cells were used. Mapping to unique ENSG ID was available for 16,885 genes. Cell types with blue bars are not significant after correction for multiple testing within or across categories. For details on different cell types and their abbreviations please refer to **Supplemental Figure S9**.

**Supplemental Figure S12: Single cell enrichment analysis for epigenomic peaks.**

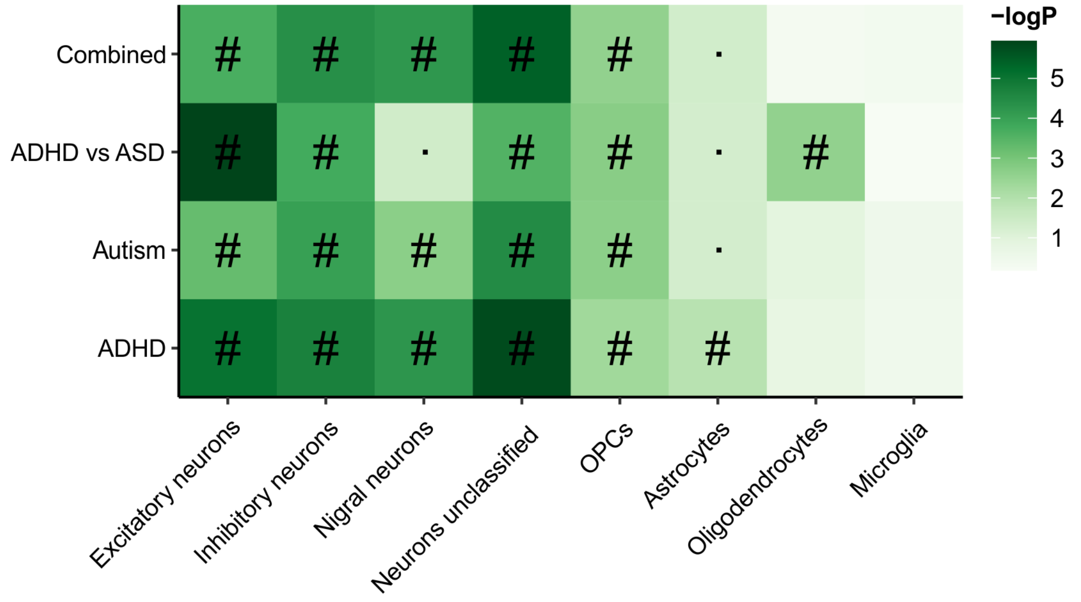

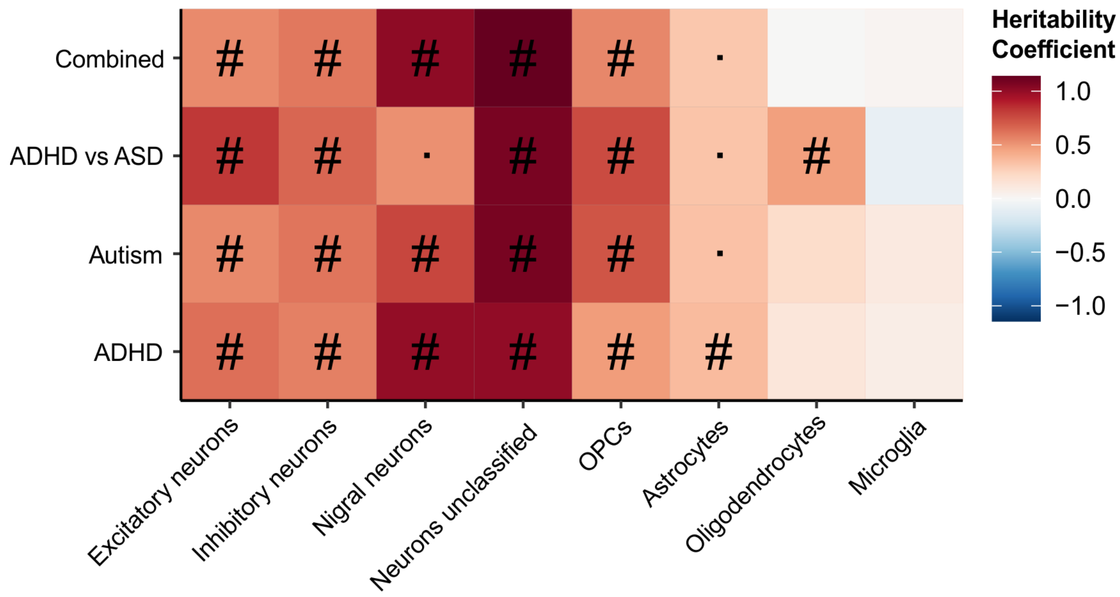

Enrichment of heritability within cell-specific open chromatin identified by scATAC-seq assay (single-cell assay for transposase accessible chromatin) calculated using LD-score partitioned heritability. Top part of figure: # - Test wide significant at FDR < 0.05; · - Nominally significant at p < 0.05. Bottom part of figure: The heritability coefficient is the regression coefficient normalized by the per-SNP heritability.

#### Supplemental Figure S13: Multivariate PRS analyses for Neuroticism subitems

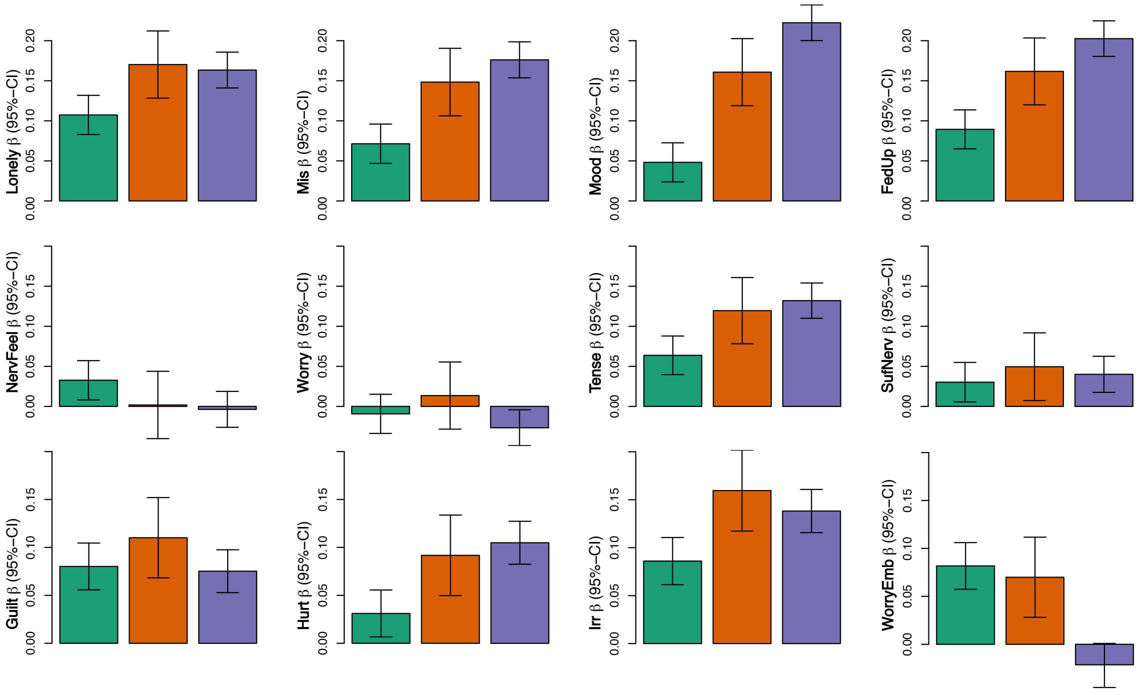

Comparison of PRS profiles across ADHD/ASD subtypes for 12 neuroticism subitems. Green bars represent ASD only cases, orange bars depict comorbid samples, and purple bars show average PRS for ADHD only cases. **Lonely** - Do you often feel lonely? (yes/no); **Mis** - Do you ever feel 'just miserable' for no reason? (yes/no); **Mood** - Does your mood often go up and down? (yes/no); **FedUp** - Do you often feel 'fed-up'? (yes/no); ***NervFeel*** - Would you call yourself a nervous person? (yes/no); ***Worry*** - Are you a worrier? (yes/no); ***Tense*** - Would you call yourself tense or 'highly strung'? (yes/no); ***SufNerv*** - Do you suffer from 'nerves'? (yes/no); **Guilt** - Are you often troubled by feelings of guilt? (yes/no); **Hurt** - Are your feelings easily hurt? (yes/no); **Irr** - Are you an irritable person? (yes/no); **WorryEmb** - Do you worry too long after an embarrassing experience? (yes/no). In the text above an underlined item (first row of traits in figure) belongs to the depressed affect cluster in Nagel et al [9] while an item in italic (second row in figure) belongs to the worry cluster. Items that are neither underlined nor italic (last row in figure) do not belong to the two clusters.

**Supplemental Figure S14: GCTA-based heritability estimates and genetic correlation for ASD (with subtypes) and ADHD**

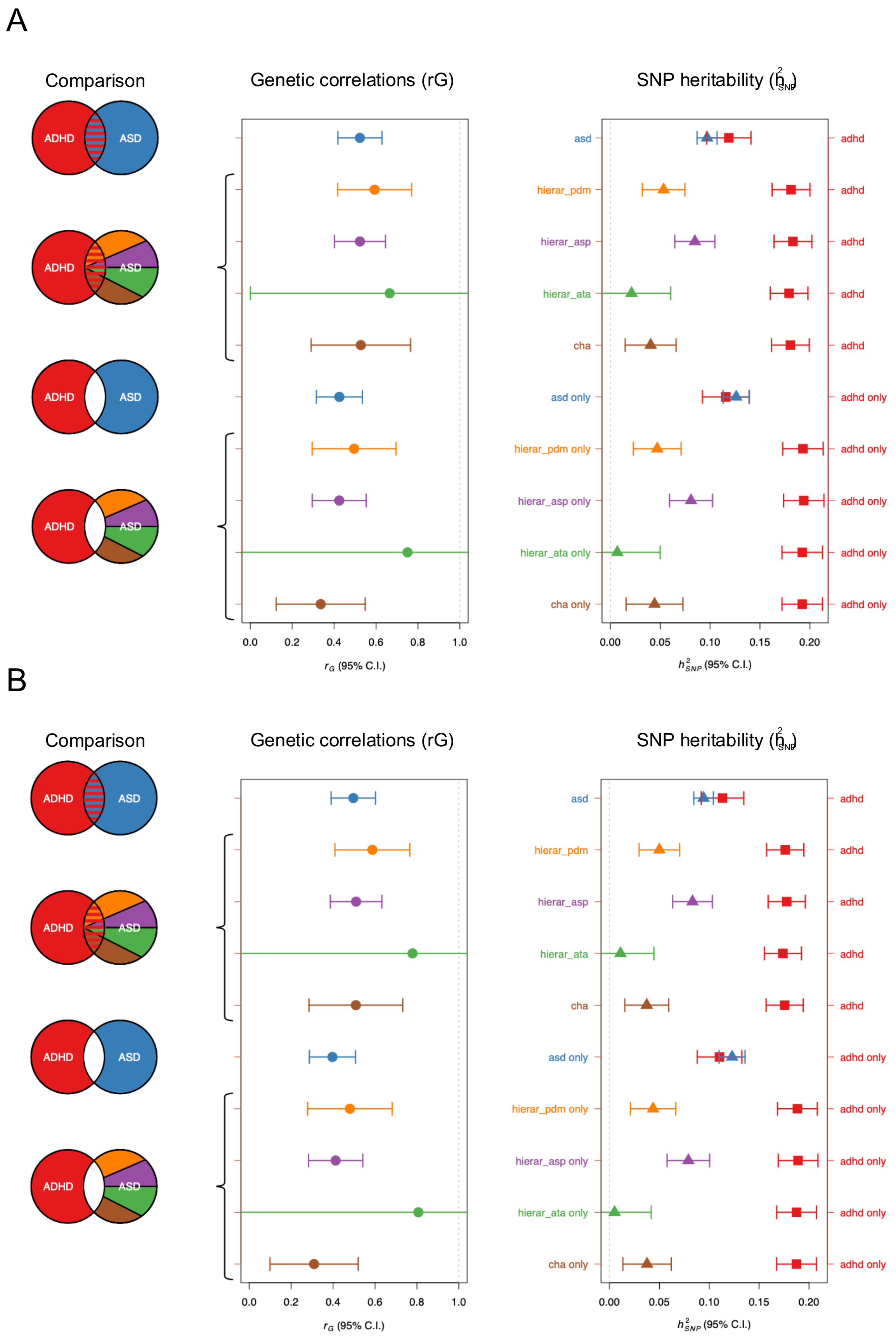

All analyses used the GCTA framework. For analyses datasets were split to allow for comparisons of independent datasets. Sample split was kept the same across analyses (with control samples using intra case ratios for splitting). For ASD subtypes a hierarchical approach was taken in the reverse order the comparison groups appear in the plot (i.e. first all individuals with childhood autism (cha), then those with atypical autism (ata) and no comorbid childhood autism, then those with Asperger's syndrome (asp) and no comorbid childhood autism or atypical, and finally the remaining individuals in the pervasive disorders group). Comparisons include **(A)** a base dataset that *excludes* individuals with mild and moderate intellectual disability and **(B)** a base dataset that *includes* individuals with mild and moderate intellectual disability. For all comparisons the following color coding applies: **red** – ADHD (i.e. without individuals with a comorbid ASD), **blue** – ASD (i.e. without individuals with comorbid ADHD), **brown** – childhood autism (*cha*, ICD10 F84.0), **green** - atypical autism (*ata*, ICD10 F84.1), **purple** – Asperger's syndrome (*asp*, ICD10 F84.5), and **orange** – pervasive disorders, unspecified and others (*pdm*, ICD10 F84.8+9). If a "only" follows the name of the group only non-comorbid cases between ADHD and ASD are includes (e.g. all ADHD cases that are not comorbid ASD cases). Please also see **Supplemental Table S7** at the end of this document.

#### Supplemental Figure S15: QQ plots for combined and ADHD vs ASD GWASs

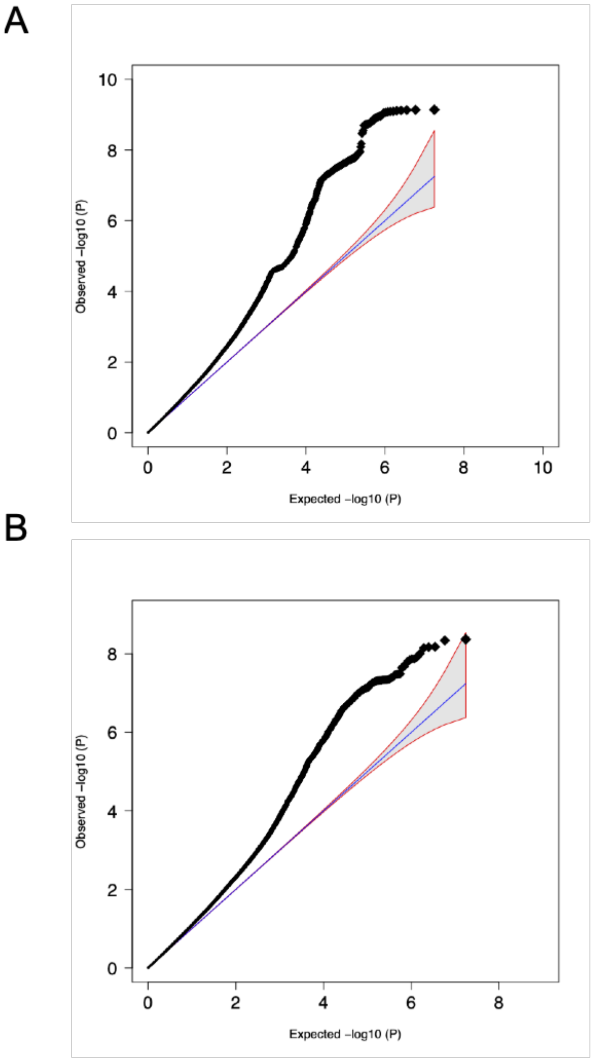

QQ-plot for **(A)** combined GWAS (34,462 cases and 41,201 controls) and for **(B)** ADHD vs ASD GWAS (11,964 ADHD only cases and 9,315 ASD only cases). The expected -log(10) under the null is plotted against the observed -log10(P) of the two aforementioned GWASs. The shading indicates 95%-confidence region under the null. The genomic inflation factor is 1.134 (with an intersect of 1.0134 in the LD score analysis) and 1.089 (intersect 0.9863) for the combined and ADHD vs ASD GWASs, respectively.
