## Supplementary material for "Identification of shared and differentiating genetic risk for autism spectrum disorder, attention deficit hyperactivity disorder and case subgroups": Table S7

### Supplemental Table S7: Genetic correlations for autism subtypes with ADHD

a)

| Trait 1 | prev(T1) | Trait 2 | prev(T2) | rG | s.e. | z | p | n |
|---|---|---|---|---|---|---|---|---|
| adhd | 0.0500 | asd | 0.0033 | 0.5231 | 0.0537 | 9.7431 | $6.54 \cdot 10^{-21}$ | 43711 |
| adhd | 0.0500 | pdm | 0.0033 | 0.5934 | 0.0900 | 6.5898 | $1.27 \cdot 10^{-10}$ | 37097 |
| adhd | 0.0500 | asp | 0.0039 | 0.5233 | 0.0622 | 8.4089 | $4.04 \cdot 10^{-16}$ | 37928 |
| adhd | 0.0500 | ata | 0.0014 | 0.6651 | 0.3392 | 1.9607 | $4.95 \cdot 10^{-02}$ | 35357 |
| adhd | 0.0500 | cha | 0.0033 | 0.5275 | 0.1211 | 4.3575 | $1.63 \cdot 10^{-05}$ | 36518 |
| adhd only | 0.0500 | asd only | 0.0100 | 0.4250 | 0.0560 | 7.5862 | $1.74 \cdot 10^{-13}$ | 41243 |
| adhd only | 0.0500 | pdm only | 0.0036 | 0.4952 | 0.1021 | 4.8514 | $1.73 \cdot 10^{-06}$ | 34626 |
| adhd only | 0.0500 | asp only | 0.0039 | 0.4243 | 0.0656 | 6.4728 | $2.60 \cdot 10^{-10}$ | 35457 |
| adhd only | 0.0500 | ata only | 0.0014 | 0.7503 | 1.1988 | 0.6259 | $5.42 \cdot 10^{-01}$ | 32886 |
| adhd only | 0.0500 | cha only | 0.0033 | 0.3360 | 0.1085 | 3.0977 | $2.00 \cdot 10^{-03}$ | 34047 |

b)

| Trait 1 | prev(T1) | Trait 2 | prev(T2) | rG | s.e. | z | p | n |
|---|---|---|---|---|---|---|---|---|
| adhd | 0.0500 | asd | 0.0033 | 0.4965 | 0.0540 | 9.1866 | $7.80 \cdot 10^{-19}$ | 45289 |
| adhd | 0.0500 | pdm | 0.0033 | 0.5874 | 0.0911 | 6.4464 | $3.05 \cdot 10^{-10}$ | 37939 |
| adhd | 0.0500 | asp | 0.0039 | 0.5099 | 0.0628 | 8.1191 | $3.65 \cdot 10^{-15}$ | 38619 |
| adhd | 0.0500 | ata | 0.0014 | 0.7791 | 0.6190 | 1.2587 | $2.10 \cdot 10^{-01}$ | 36221 |
| adhd | 0.0500 | cha | 0.0033 | 0.5087 | 0.1143 | 4.4522 | $1.08 \cdot 10^{-05}$ | 37682 |
| adhd only | 0.0500 | asd only | 0.0100 | 0.3968 | 0.0562 | 7.0602 | $6.25 \cdot 10^{-12}$ | 42619 |
| adhd only | 0.0500 | pdm only | 0.0036 | 0.4802 | 0.1030 | 4.6633 | $4.16 \cdot 10^{-06}$ | 35265 |
| adhd only | 0.0500 | asp only | 0.0039 | 0.4122 | 0.0662 | 6.2263 | $1.14 \cdot 10^{-09}$ | 35945 |
| adhd only | 0.0500 | ata only | 0.0014 | 0.8068 | 1.4946 | 0.5398 | $6.02 \cdot 10^{-01}$ | 33547 |
| adhd only | 0.0500 | cha only | 0.0033 | 0.3092 | 0.1074 | 2.8791 | $4.04 \cdot 10^{-03}$ | 35008 |

All analyses used the GCTA [10] framework. For analyses datasets were split to allow for comparisons of independent datasets. Sample split was kept the same across analyses (with control samples using intra case ratios for splitting). For ASD subtypes a hierarchical approach was taken: ***cha*** – Childhood autism (ICD10 F84.0), ***ata*** – Atypical autism (ICD10 F84.1), without comorbid CHA, ***asp*** – Asperger's syndrome (ICD10 F84.5), without comorbid CHA and/or ATA, ***pdm*** – Pervasive disorders, unspecified and other, ICD10 F84.8+9), without any of the previous. Comparisons include **(a)** a base dataset that *excludes* individuals with mild and moderate intellectual disability and **(b)** a base dataset that *includes* individuals with mild and moderate intellectual disability. *Trait 1 and 2* – traits in the comparison (an "only" in the name of the traits means removal of comorbid ADHD and ASD, respectively, e.g. ADHD only – individuals ADHD and without ASD diagnose. For ASD subtypes the comorbid ADHD cases are removed); ***prev(T1) / prev(T2)*** – population prevalence for traits 1 and 2, respectively; ***$r_G$*** – genetic correlation; ***s.e.*** – standard error; ***z*** – Z score; ***p*** – p-value for genetic correlation using an approach by Altman and Bland (doi: 10.1136/bmj.d2304); ***n*** – number of samples in analysis.
